# Modelling the health and economic impacts of M72/AS01_E_ vaccination and BCG-revaccination: estimates for South Africa

**DOI:** 10.1101/2023.10.04.23296538

**Authors:** Tom Sumner, Rebecca A. Clark, Christinah Mukandavire, Allison Portnoy, Chathika K. Weerasuriya, Roel Bakker, Danny Scarponi, Mark Hatherill, Nicolas A. Menzies, Richard G. White

## Abstract

**Background:** Tuberculosis remains a major public health problem in South Africa, with an estimated 300,000 cases and 55,000 deaths in 2021. New tuberculosis vaccines could play an important role in reducing this burden. Phase IIb trials have suggested efficacy of the M72/AS01_E_ vaccine candidate and BCG-revaccination. The potential population impact of these vaccines is unknown.

**Methods:** We used an age-stratified transmission model of tuberculosis, calibrated to epidemiological data from South Africa, to estimate the potential health and economic impact of M72/AS01_E_ vaccination and BCG-revaccination. We simulated vaccination scenarios over the period 2025–2050 with a range of product characteristics and delivery strategies. We calculated reductions in tuberculosis cases and deaths and costs and cost-effectiveness from health-system and societal perspectives.

**Results:** M72/AS01_E_ vaccination may have a larger impact than BCG-revaccination, averting approximately 80% more cases and deaths by 2050. Both vaccines were found to be cost-effective (compared to no new vaccine) across a range of vaccine characteristics and delivery strategies. The impact of M72/AS01_E_ is dependent on the assumed efficacy of the vaccine in uninfected individuals. Extending BCG-revaccination to HIV-infected individuals on ART had minimal effect on the health impact, but increased costs by approximately 70%.

**Conclusions:** Our results show that M72/AS01_E_ vaccination or BCG-revaccination could be cost-effective in South Africa. However, there is considerable uncertainty in the estimated impact and costs due to uncertainty in vaccine characteristics and the choice of delivery strategy.

## Introduction

Tuberculosis (TB) continues to be a major cause of illness and mortality with an estimated 10.6 million incident cases and 1.6 million deaths in 2021 [1]. While TB incidence and mortality had been declining prior to the COVID pandemic, the rate of decline is insufficient to meet the Sustainable Development Goals target to end the tuberculosis epidemic by 2030 [4]. Previous work [2, 3] has shown that new TB vaccines will be critical to accelerate declines in TB and achieve elimination.

Several new vaccine candidates are in development [5]. Candidate M72/AS01_E_ has been shown to have efficacy of 49.7% (95% confidence interval: 2.1–74.2) against TB disease over 3 years follow-up among adolescents and adults with previous sensitisation to *Mycobacterium tuberculosis* (*M.tb*), indicated by a positive QuantiFERON-TB Gold In-tube assay (QFT) result [6]. There is also renewed interest in the use of Bacillus Calmette-Guérin (BCG) revaccination to prevent TB. Evaluation of BCG-revaccination of QFT-negative adolescents showed an efficacy of 45.5% (6.4–68.1) against the secondary endpoint, sustained QFT conversion at 6 months [7].

Recent modelling [8] has suggested that M72/AS01_E_ vaccination and BCG-revaccination could be cost-effective in India for a range of different vaccine characteristics and implementation strategies. However, the potential impact and costs of vaccine implementation are likely to be context-specific. Previous analyses of the impact of hypothetical TB vaccines in China, South Africa, and India showed that overall vaccine impact, and the importance of different vaccine characteristics, was dependent on the epidemiology in each country [3].

TB remains a major public health problem in South Africa. Despite huge improvements in antiretroviral therapy (ART) coverage, human immunodeficiency virus (HIV) continues to be a significant factor in the TB epidemic. In 2021, over 50% of incident TB cases and almost 60% of deaths due to TB were among people living with HIV (PLHIV) [1]. Previous modelling of routine adolescent vaccination [9] or mass vaccination of adults [10] with an M72/AS01_E_-like vaccine found it was likely to be cost-effective in South Africa, but the cost-effectiveness of other implementation strategies for M72/AS01_E_ vaccination or BCG-revaccination in this setting have not been explored. In this paper, we used a mathematical model to estimate the health impact and cost-effectiveness of M72/AS01_E_ and BCG-revaccination in South Africa under a range of vaccine characteristics and delivery assumptions.

## Methods

### Data

We used estimates for South Africa’s demography from the United Nations Population Division [11]. TB incidence and mortality estimates and reported notifications were taken from the WHO [1] and prevalence data were obtained from the South African national prevalence survey [12]. HIV incidence, prevalence, and mortality estimates and data on ART coverage and levels of viral suppression were obtained from UNAIDS [13]. TB and HIV data values and sources are reported in the supplementary material (section 3).

### Model

We developed a mathematical model of TB in South Africa. The model is an adaptation of previous models of TB vaccination [8, 14]. It describes infection with *M.tb*, progression of TB disease, and diagnosis and treatment, stratified by access to care, age, and vaccination status.

HIV is a key risk factor for TB in South Africa. We stratified the model by HIV status to capture the stages of HIV infection and treatment relevant for TB epidemiology and to simulate differences in the targeting and efficacy of TB vaccines by HIV status. The HIV-infected population was stratified by CD4 count (>350 cells per cubic millimeter, <350 cells per cubic millimeter), whether individuals had been diagnosed with HIV or not, whether they are on ART or not, and whether they are virally suppressed or not.

Full details of the model structure and parameter values are given in the supplementary material (section 1 and 2).

### Calibration

The model was fitted to TB and HIV calibration targets using history matching with emulation, a method that allows efficient exploration of high-dimensional parameter spaces and generates a reduced parameter space that is consistent with the calibration targets [15]. History matching was implemented using the *hmer* package in R [16]. Further details of the process are given in the supplementary material (section 3).

Calibration targets included: TB incidence rate by HIV status and age; TB mortality rate by HIV status; TB notification rate; overall TB prevalence; the proportion of TB prevalence that was sub-clinical (asymptomatic); the ratio of prevalent TB in low and high access to care strata; HIV prevalence by age; HIV (AIDS-related) mortality; the proportion of PLHIV who knew their status. Values and sources for the calibration targets are given in table S3.1 in the supplementary material.

TB diagnosis and treatment were explicitly included in the model but existing prevention measures (neonatal BCG vaccination and preventive therapy) were not. We assumed that the effects of these prevention measures are included in the calibration targets and therefore implicitly captured in our model.

### Vaccine scenarios

In our baseline (no-new-vaccine) scenario, we assumed that TB and HIV diagnosis and treatment continued at 2020 levels, and that HIV incidence remained constant at 2020 estimates. The calibrated model was used to simulate the TB and HIV epidemiology to 2050 under this no-new-vaccine scenario.

The basecase M72/AS01_E_ vaccination scenario assumed a vaccine with 50% efficacy against TB disease [6] and 10-year duration of protection [17]. The efficacy of M72/AS01_E_ has only been measured in QFT-positive individuals [6]; however, immunogenicity studies suggest that two-doses of M72/AS01_E_ induced sustained antigen-specific T cell and IgG responses in both QFT-positive and QFT-negative adolescents [18]. In our main analysis, we therefore assumed the vaccine would be efficacious irrespective of *M.tb* infection status at time of vaccination. We also explored scenarios in which M72/AS01_E_ was only efficacious in individuals with *M.tb* infection. We simulated the introduction of the vaccine in 2030 with routine vaccination of 15-year-olds (80% coverage) and campaigns for 16–34-year-olds conducted in 2030 and 2040 (70% coverage). We assumed that M72/AS01_E_ would be given to all individuals irrespective of HIV status with reduced efficacy in PLHIV [19] (10% reduction in efficacy in virally suppressed individuals; 46% reduction in efficacy in all other PLHIV) (see supplementary material section 4). We assumed that a course of M72/AS01_E_ vaccination would require two doses, with a price of US$2.50 per dose.

The basecase BCG-revaccination scenario assumed a vaccine with 45% efficacy against sustained *M.tb* infection [7] with 10-year duration of protection [17]. We assumed the vaccine would only be efficacious in individuals uninfected with *M.tb* at time of vaccination. We simulated the introduction of revaccination in 2025, with routine vaccination of 10-year-olds (80% coverage) and campaigns for 11–18-year-olds in 2025, 2035, and 2045 (80% coverage). Due to uncertainty about the safety of BCG in PLHIV (especially infants) [20], in the basecase scenario we assumed that BCG-revaccination would only be offered to HIV-uninfected individuals. We assumed that BCG-revaccination would require a single dose at the average UNICEF BCG price of US$0.17 per dose [21].

Vaccine introduction costs for both vaccine products were assumed to be US$2.40 (range: 1.20–4.80) per individual in the targeted age group based on vaccine introduction support policy from Gavi, the Vaccine Alliance [22]. A further US$0.11 (0.06–0.22) supply costs and US$2.50 (1.00–5.00) delivery costs per dose were included [23]. We assumed vaccine a wastage rate of 5%.

### Uncertainty analysis

We considered several different delivery options for M72/AS01_E_ vaccination and BCG-revaccination (Table 1). These represented a finite set of discrete policies for vaccine delivery that may be considered by decision makers. For M72/AS01_E_ vaccination, we considered policies based on different age targeting. For BCG-revaccination, we considered different age targeting and, based on evidence that the protective benefit of BCG can outweigh the risks of local or disseminated BCG disease in people established on ART [24], policies including revaccination of PLHIV who are on ART and/or virally suppressed.

**Table 1.**
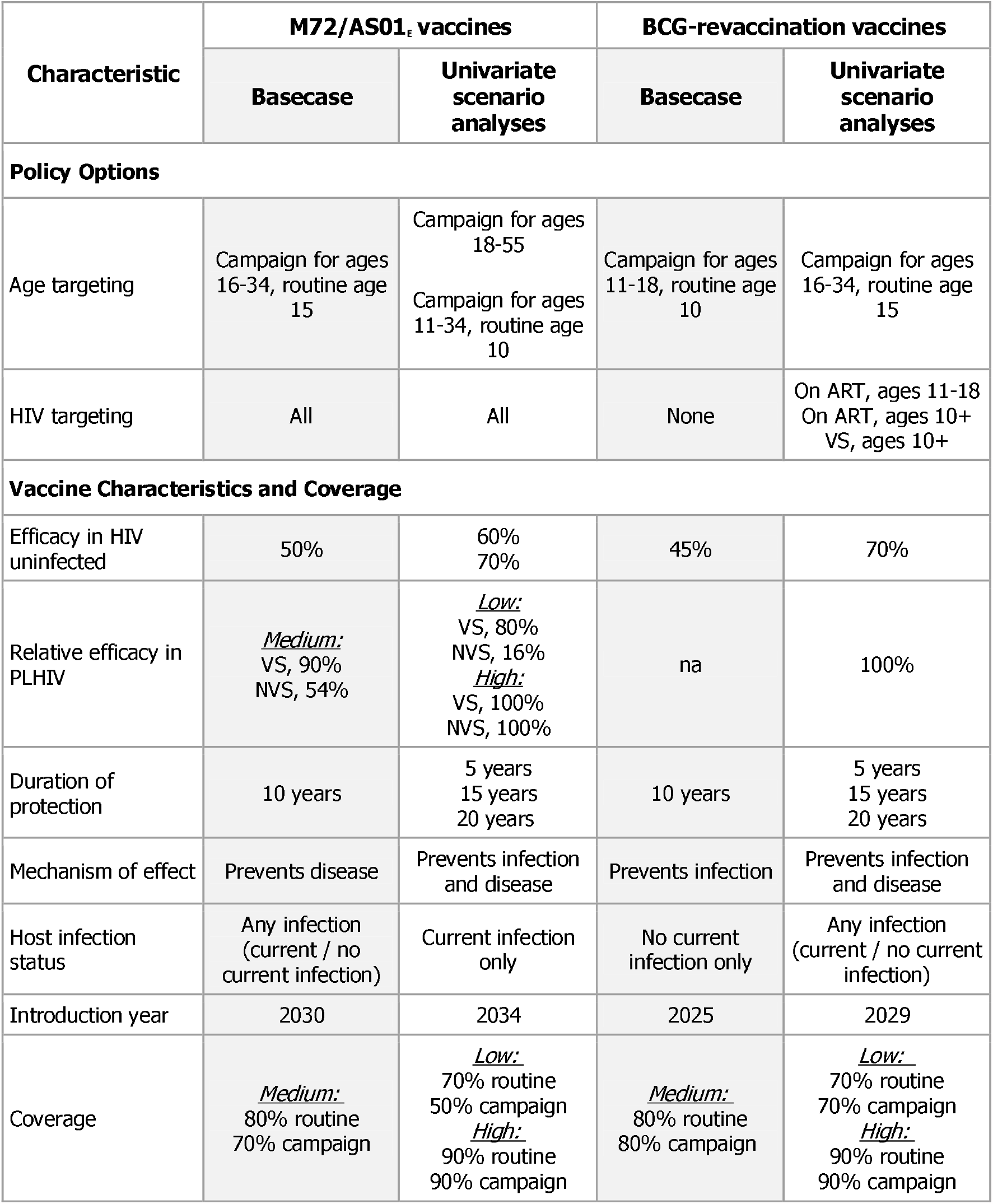
Vaccine scenarios. Basecase assumptions are shown in the shaded columns. Other assumptions are explored in univariate scenario analysis. VS = virally suppressed, NVS = not virally suppressed

We explored a variety of different vaccine characteristics to explore the effects of uncertainty on the results (Table 1). These include varying vaccine efficacy, duration of protection, year of introduction, and coverage. We explored scenarios in which M72/AS01_E_ vaccination and BCG-revaccination were efficacious against both sustained *M.tb* infection and TB disease, and scenarios in which M72/AS01_E_ efficacy was limited to individuals who were infected with *M.tb* at time of vaccination and BCG-revaccination was efficacious irrespective of *M.tb* infection status at time of vaccination. For M72/AS01_E_ vaccination, we explored two different assumptions about the efficacy in PLHIV.

### Outcomes

We calculated the cumulative number of TB cases and deaths averted by 2050 compared to the no-new-vaccine baseline. We estimated the annual incremental costs over the same timeframe in 2020 US dollars from both health-system and societal perspectives. Health-system costs included vaccine costs, costs of testing and treating TB, and the costs of ART. The societal perspective also included costs of patient time for vaccination, non-medical patient costs (e.g., transportation) associated with TB, and indirect patient costs of TB treatment. Full details of the costs are in table S5.1 in the supplementary material. Total disability-adjusted life years (DALYs) averted from the introduction of vaccination to 2050 were calculated using disability weights for TB disease, and aspirational life tables, from the Global Burden of Disease 2019 study [25].

Cost-effectiveness analysis was conducted for the policy options for each vaccine. The difference in incremental cost-effectiveness ratios (ICERs) (the ratio of mean incremental costs to mean incremental benefits in DALYs averted) were calculated from a health-system perspective for each non-dominated strategy for the analytic period 2025–2050. We compared the cost-effectiveness estimates to three country specific thresholds: 1x gross domestic product (GDP) per capita in 2020 (US$5,742) [26] and the upper and lower bounds of country-level opportunity cost thresholds defined by Ochalek et al. [27] (Ochalek upper [US$3,334] and lower [US$2,480] bounds).

To explore how cost-effectiveness of vaccination depended on vaccine characteristics and coverage assumptions we examined the difference in ICERs for the characteristics listed in Table 1, compared to the no-new-vaccine baseline, assuming the vaccine was introduced using the basecase policy option. ICERs were calculated from a health system and societal perspective.

## Results

### No-new-vaccine projection

In the absence of new vaccines, there were an estimated 9.4 (95% uncertainty interval: 8.6– 11.0) million incident TB cases and 1.6 (1.4–1.9) million deaths due to TB between 2025 and 2050. Plots of the baseline model outputs can be found in the supplementary material (section 7).

### Health impact

With our basecase assumptions, M72/AS01_E_ vaccination could avert 1.56 (1.44–1.87) million cases and 0.22 (0.18–0.25) million deaths by 2050 (Figure 1). If vaccine efficacy was higher (70% vs. 50%), the number of cases averted could be 33% higher. If the vaccine protected against both disease and infection, the impact could be 37% higher. When we assumed that M72/AS01_E_ was only effective in people who were infected with *M.tb* at the time of vaccination, the number of cases averted was 40% lower. The relative efficacy of M72/AS01_E_ in PLHIV compared to HIV-uninfected individuals produced a relatively small change in the predicted impact. If the vaccine was equally efficacious in both populations, the numbers of cases and deaths averted could be increased by 7% and 10%, respectively.

**Figure 1.**
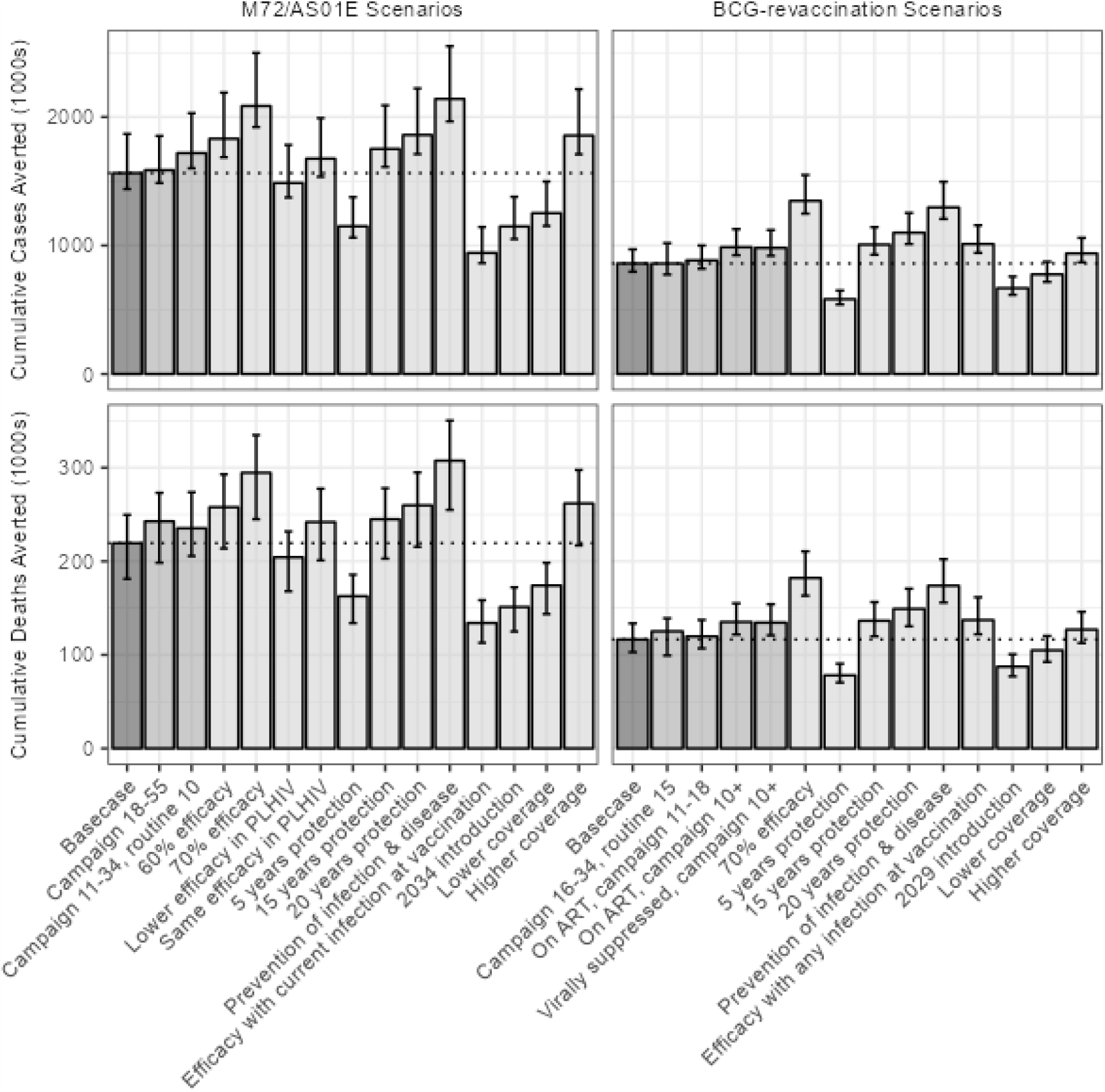
Cumulative cases and deaths averted by 2050. Bars show the median estimates and error bars show the 95% uncertainty range. The horizontal dashed line indicates the median estimate from the Basecase scenario. Shading indicates the scenario types (dark grey: Basecase; mid grey: policy options; light grey: vaccine characteristics and coverage).

In the basecase, BCG-revaccination could avert 0.86 (0.80–0.97) million cases and 0.12 (0.10–0.13) million deaths by 2050 (Figure 1). Similar to M72/AS01_E_, increased efficacy (70% vs. 45%) could increase the number of cases averted by 57%, while assuming protection against infection and disease could increase the impact by 51%. Delayed introduction of BCG-revaccination (by 4 years to 2029) resulted in a reduction of 22% in the number of cases averted. We found a small additional impact (3% increase in cases averted) when extending BCG-revaccination to 11–18-year-olds on ART (compared to no use of BCG in PLHIV) and 15% greater impact if BCG-revaccination was offered to all people on ART aged 10 years or older.

In our basecase scenarios, we found an 82% greater impact from M72/AS01_E_ vaccination compared to BCG-revaccination (1.56 million cases averted vs 0.86 million cases averted). If M72/AS01_E_ was assumed to only be effective in people who were infected with *M.tb* at the time of vaccination then the number of cases and deaths averted was comparable to that predicted for the basecase BCG-revaccination scenario.

### Cost-effectiveness

Compared to the no-new-vaccine scenario, the basecase M72/AS01_E_ policy had an incremental cost of US$255 million from a health system perspective, US$64 million from a societal perspective and averted 3.7 million DALYs by 2050. A policy which vaccinated older age groups (campaign for ages 18–55) was dominated by the other policy options considered (i.e., this strategy was more costly and less effective than alternative strategies) and was removed from further analysis (Table 2). The basecase (routine vaccination for age 15, campaign for ages 16–34,) and a policy of delivering the vaccine to younger ages (routine vaccination for age 10, campaign for ages 11–34,) were found to be potentially cost-effective from a health system perspective and are displayed on the efficiency frontier in Figure 2.

**Table 2.**
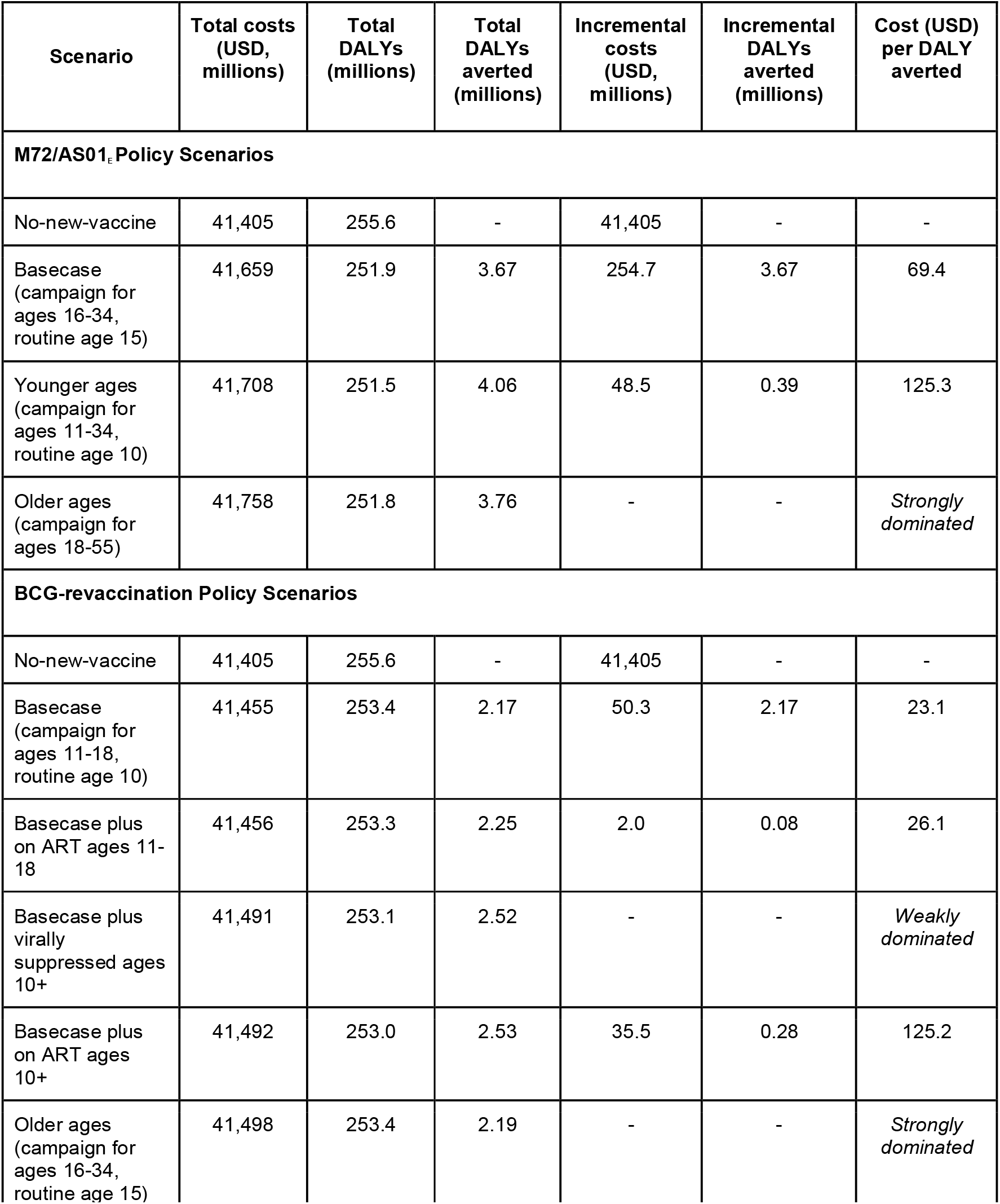
Cost-effectiveness analysis of policy scenarios. Discounted costs from a health-system perspective (US$ millions). Discounted (DALYs) (millions)

| Scenario | Total costs<br>(USD,<br>millions) | Total<br>DALYs<br>(millions) | Total<br>DALYs<br>averted<br>(millions) | Incremental<br>costs<br>(USD,<br>millions) | Incremental<br>DALYs<br>averted<br>(millions) | Cost (USD)<br>per DALY<br>averted |
| --- | --- | --- | --- | --- | --- | --- |
| **M72/AS01<sub>E</sub> Policy Scenarios** |  |  |  |  |  |  |
| No-new-vaccine | 41,405 | 255.6 | - | 41,405 | - | - |
| Basecase<br>(campaign for<br>ages 16-34,<br>routine age 15) | 41,659 | 251.9 | 3.67 | 254.7 | 3.67 | 69.4 |
| Younger ages<br>(campaign for<br>ages 11-34,<br>routine age 10) | 41,708 | 251.5 | 4.06 | 48.5 | 0.39 | 125.3 |
| Older ages<br>(campaign for<br>ages 18-55) | 41,758 | 251.8 | 3.76 | - | - | *Strongly*<br>*dominated* |
| **BCG-revaccination Policy Scenarios** |  |  |  |  |  |  |
| No-new-vaccine | 41,405 | 255.6 | - | 41,405 | - | - |
| Basecase<br>(campaign for<br>ages 11-18,<br>routine age 10) | 41,455 | 253.4 | 2.17 | 50.3 | 2.17 | 23.1 |
| Basecase plus<br>on ART ages 11-18 | 41,456 | 253.3 | 2.25 | 2.0 | 0.08 | 26.1 |
| Basecase plus<br>virally<br>suppressed ages<br>10+ | 41,491 | 253.1 | 2.52 | - | - | *Weakly*<br>*dominated* |
| Basecase plus<br>on ART ages<br>10+ | 41,492 | 253.0 | 2.53 | 35.5 | 0.28 | 125.2 |
| Older ages<br>(campaign for<br>ages 16-34,<br>routine age 15) | 41,498 | 253.4 | 2.19 | - | - | *Strongly*<br>*dominated* |

**Figure 2.**
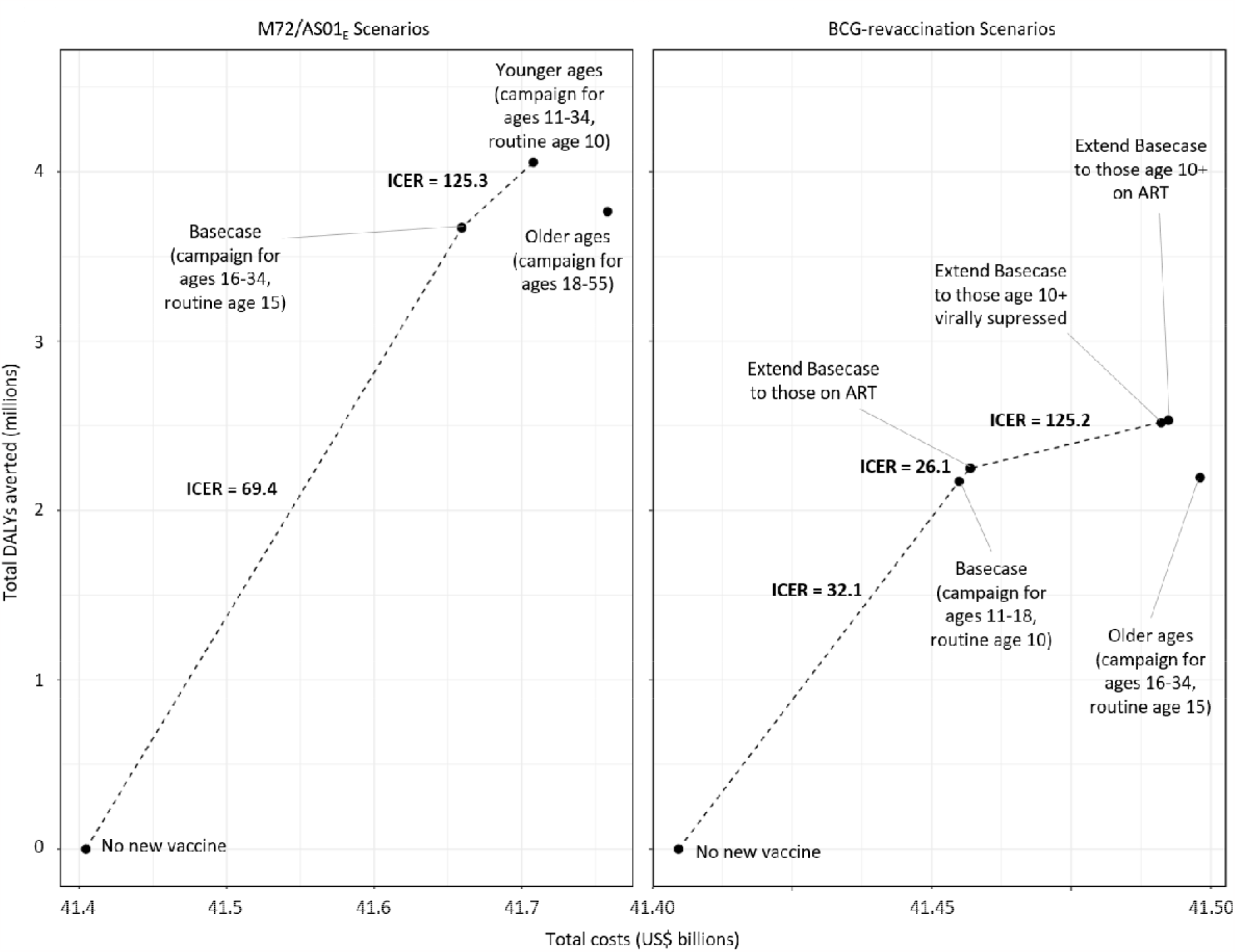
Efficiency frontiers for policy options. Discounted total costs (US$ billions) from a health-system perspective versus discounted total disability-adjusted life years (DALYs) averted (millions) for each policy option. Dashed lines indicate the efficiency frontiers. Incremental cost effectiveness ratios (ICERs) are shown in US$ per DALY averted.

The basecase BCG-revaccination policy had an incremental cost of US$50 million (health system), was cost-saving from a societal perspective and averted 2.2 million DALYs by 2050 compared to the no-new-vaccine baseline. For BCG-revaccination, of the policies modelled, those which targeted older ages (routine age 15, campaign for ages 16–34) or included virally suppressed individuals aged over 10 years were both dominated by other strategies and removed from consideration. Including individuals on ART aged 10 years or over was the optimal option of the strategies considered.

Figure 3 shows the ICERs (health-system perspective) for the scenarios with different vaccine characteristics and coverages, assuming that the vaccines are introduced using the basecase policy option (results for the societal perspective are presented in supplementary material section 9). Our results suggest that the introduction of M72/AS01_E_ would be cost-effective compared to not introducing the vaccine irrespective of vaccine characteristics. BCG-revaccination would be cost-effective from a health system perspective or cost-saving from a societal perspective.

**Figure 3.**
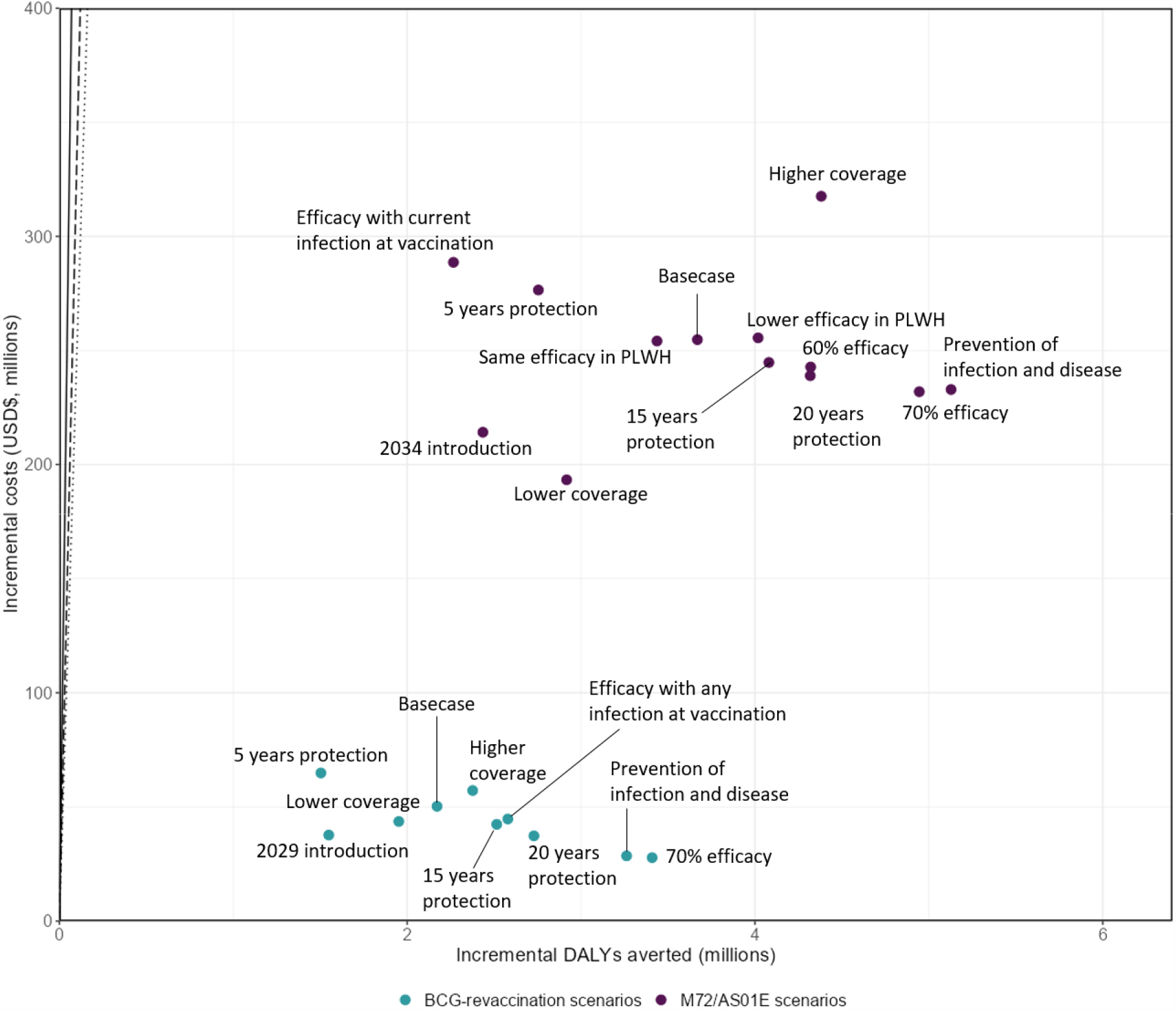
Comparison of ICERs for varying vaccine characteristics and coverage. Discounted incremental disability-adjusted life years (DALY) averted (millions) versus discounted incremental costs (US$ millions) for each scenario compared to the no-new-vaccine baseline. Points show the mean values for each characteristic, assuming vaccines are introduced using the Basecase policy option. Lines (on far left) indicate cost-effectiveness thresholds based on 1x per-capita GDP (solid line), the Ochalek upper bound (dashed line), and the Ochalek lower bound (dotted line). Points lying to the right of a given line indicate that the scenario would be considered cost-effective compared to the no-new-vaccine baseline.

In the basecase policy scenario, the costs of vaccinating with M72/AS01_E_ were approximately US$12 million per year compared to approximately US$3 million for BCG-revaccination, in part driven by the lower cost of BCG. The incremental costs of the basecase M72/AS01_E_ scenario were approximately five times higher than those for BCG-revaccination, due to larger increases in future ART costs associated with the reductions in HIV-TB mortality. From a societal perspective, both vaccines resulted in reductions in direct and indirect patient costs due to the reduced burden of TB (see supplementary material section 9 for breakdown of the costs of each scenario).

## Discussion

Our results suggest that, across the scenarios considered, M72/AS01_E_ vaccination in South Africa could avert between 0.86 and 2.55 million TB cases and 0.11 to 0.35 million deaths by 2050. BCG-revaccination could avert 0.54 to 1.55 million cases and 0.07 to 0.17 million deaths. M72/AS01_E_ vaccination had higher incremental costs than BCG-revaccination due to the higher costs of M72/AS01_E_ resulting in higher ICERs for M72/AS01_E_. All scenarios we simulated were cost-effective at all thresholds considered under a health system perspective, and cost-effective or cost-saving under a societal perspective, consistent with previous cost-effectiveness estimates of novel TB vaccination in other settings [8, 28].

We used data from clinical trials [6, 7] to inform our basecase vaccine characteristics. However, several important characteristics, such as the duration of protection, are unknown. Similarly, our policy scenarios were informed by expert opinion, but the real-world implementation will also be dependent on logistical and operational criteria. To address this, we explored a variety of vaccine characteristics and delivery scenarios. As expected, increasing vaccine efficacy, duration of protection, vaccination coverage, and the age range of the population offered vaccination increased the estimated impact. Based on clinical trial endpoints [6, 7], we assumed in our basecase scenarios that M72/AS01_E_ provided protection against progression to disease while BCG-revaccination provided protection against sustained infection with *M.tb*. These assumptions are important in determining the results, with the addition of protection against infection or disease for M72/AS01_E_ or BCG, respectively, significantly increasing the predicted impact and reducing the ICERs. The host status required for the vaccine to be effective was also important for M72/AS01_E_ vaccination. We assumed, based on immunogenicity studies [18], that the vaccine would be effective in all individuals whether they had previously been infected with *M.tb* or not. However, to date, efficacy of M72/AS01_E_ has only been directly assessed in QFT-positive individuals [6]. When we restricted the efficacy in our model to currently infected individuals, the impact of M72/AS01_E_ was reduced to similar levels as BCG-revaccination and the ICERs approximately doubled. Data on the efficacy of M72/AS01_E_ in QFT-negative individuals, together with improved estimates of QFT positivity rates in target populations, will be important for refining estimates of the impact and cost-effectiveness of M72/AS01_E_ vaccination.

The potential effect of M72/AS01_E_ in HIV-infected individuals is unclear. While safety and immunogenicity trials have shown it is likely to be safe and immunogenic in this population [19], there is no clinical trial evidence of efficacy against TB disease or *M.tb* infection. We considered three scenarios for the efficacy of M72/AS01_E_ in HIV-infected people and found that this assumption had only a small effect on the predicted impact. This is in part due to the relatively small population affected (compared to the HIV-uninfected population) and the narrow range of the relative efficacy parameter informed by immunogenicity data [19].

While BCG is contraindicated in HIV-infected infants [20], there is evidence that its protective benefit can outweigh the risks of BCG disease in people established on ART [24]. In our basecase, we assumed that BCG would be given to HIV-uninfected individuals and explored the effect of extending BCG-revaccination to people on ART in scenario analysis. Including HIV-infected individuals without extending the age range of BCG-revaccination (10–18-year-olds) had minimal effect on the results because of the small additional number of people vaccinated. When BCG-revaccination was extended to all people on ART aged 10 or over we found an approximate 15% increase in the number of cases averted. This increase was based on the assumption that BCG revaccination would have equivalent efficacy in PLHIV on ART as in HIV-uninfected individuals. If the efficacy of BCG is reduced in this population then the additional benefit would be smaller. The expansion of BCG targeting to PLHIV also increased the health system costs for BCG-revaccination. This increase in costs, from US$50 million to US$87 million, is due to the increased number of vaccines given and increases in future ART costs.

Comparison with our previous modeling of M72/AS01_E_ vaccination and BCG-revaccination in India [8] highlights some key similarities and differences between countries. In our basecase scenarios M72/AS01_E_ vaccination prevented more cases of TB than BCG-revaccination in both countries. The additional benefit of M72/AS01_E_ was greater in South Africa (80% more cases averted by M72/AS01_E_ than BCG) than in India (40% more cases averted by M72/AS01_E_ than BCG). The differences in impact between M72 and BCG are partly a result of the assumed characteristics of the vaccines and also the modeled prevalence of infection in the two countries. We assumed that BCG-revaccination was only efficacious in those who are not infected at time of vaccination and therefore the relative effect of BCG-revaccination is lower in South Africa where we assumed a higher prevalence of prior infection with *M.tb*. In India, the incremental cost of vaccine roll-out was greater than in South Africa (20-fold higher for M72/AS01_E_, 12-fold higher for BCG-revaccination) as a result of the larger population size in India. For M72/AS01_E_, adjusting vaccine characteristics had similar effects on the cost-effectiveness results in both countries. In India, the cost-effectiveness of BCG-revaccination was more sensitive to assumed vaccine efficacy and age targeting, than in South Africa.

Our results are derived from a mathematical model and are therefore subject to several limitations in addition to the uncertainty about vaccine characteristics. Our model was based on the most recent knowledge on the clinical spectrum of TB, incorporating subclinical states and declining risks of disease by time since infection. Uncertainty in these assumptions and their interactions with the efficacy of vaccines may affect our results [29]. In generating our baseline projection of future TB burden we have assumed that TB care will continue at current levels in the future. Changes in diagnostics, drugs or preventive therapy may alter this trajectory and may affect the estimated impact of new vaccines. We also made several simplifications in our modelling of HIV to ensure a workable number of states in our model, and made assumptions about the efficacy of vaccination in people living with HIV. Further work will explore in more detail the interaction between HIV, levels of immunosuppression, and vaccine efficacy.

## Conclusion

Our results suggest that M72/AS01_E_ vaccination or BCG-revaccination could be cost-effective in South Africa for a range of vaccine characteristics and delivery strategies. Greatest impact could be achieved with M72/AS01_E_ vaccination while the lowest costs were associated with BCG-revaccination. However, there is considerable uncertainty in the estimated impact and costs due to uncertainty in vaccine characteristics and choice of delivery strategy. These results can help inform global and country-level decision makers on when, where and how to employ new TB vaccines.

## Data Availability

All data used in this work is summarised in the supplementary material. All model code will be made freely available upon publication.

## Ethics approval and consent to participate

Not applicable

## Consent for publication

Not applicable

## Availability of data and materials

All data used is summarised in Additional file 1.

All model code will be made freely available upon publication.

## Competing interests

The authors declare that they have no competing interests

## Funding

We thank the Bill & Melinda Gates Foundation for providing funding (INV-001754) to undertake this research.

## Author contributions

Conception: RAC, CM, AP, CKW, NAM, RGW

Data acquisition and preparation: TS, RAC, CM, AP, CKW, RB, DS, NAM, RGW

Data analysis: TS, RAC, CM, AP, CKW, NAM, RGW

Interpretation of results: TS, RAC, CM, AP, MH, NAM, RGW

Manuscript drafting and revisions: TS, RAC, CM, AP, CKW, RB, DS, MH, NAM, RGW

All authors had the opportunity to access and verify the data and were responsible for the decision to submit the manuscript for publication.

## Notes

### Competing Interest Statement

The authors have declared no competing interest.

## References

1. World Health Organisation. Global Tuberculosis Report. 2022 [accessed 2021; Available from: https://apps.who.int/iris/handle/10665/363752.

2. Weerasuriya, C.K., et al., New tuberculosis vaccines: advances in clinical development and modelling. J Intern Med, 2020. 288(6): p. 661–681.

3. Harris, R.C., et al., Potential impact of tuberculosis vaccines in China, South Africa, and India. Sci Transl Med, 2020. 12(564).

4. United Nations Department of Economic and Social Affairs, S.D. Sustainable Development Goals. 2023 [accessed 2023; Available from: https://sdgs.un.org/goals.

5. Stop TB Partnership Working Group on New TB Vaccines. TB Vaccine Pipeline. 05/06/2023]; Available from: https://newtbvaccines.org/tb-vaccine-pipeline/.

6. Tait, D.R., et al., Final Analysis of a Trial of M72/AS01(E) Vaccine to Prevent Tuberculosis. N Engl J Med, 2019. 381(25): p. 2429–2439.

7. Nemes, E., et al., Prevention of M. tuberculosis Infection with H4:IC31 Vaccine or BCG Revaccination. N Engl J Med, 2018. 379(2): p. 138–149.

8. Clark, R.A., et al., New tuberculosis vaccines in India: Modelling the potential health and economic impacts of adolescent/adult vaccination with M72/AS01 (E) and BCG-revaccination. medRxiv, 2023.

9. Harris, R.C., et al., Cost-effectiveness of routine adolescent vaccination with an M72/AS01(E)-like tuberculosis vaccine in South Africa and India. Nat Commun, 2022. 13(1): p. 602.

10. Jayawardana, S., et al., Feasibility of novel adult tuberculosis vaccination in South Africa: a cost-effectiveness and budget impact analysis. NPJ Vaccines, 2022. 7(1): p. 138.

11. United Nations Department of Economic and Social Affairs, P.D. World Population Projections (2019 revision). 2019; Available from: https://population.un.org/wpp/Download/Standard/Population/.

12. Moyo, S., et al., Prevalence of bacteriologically confirmed pulmonary tuberculosis in South Africa, 2017-19: a multistage, cluster-based, cross-sectional survey. Lancet Infect Dis, 2022. 22(8): p. 1172–1180.

13. Joint United Nations Programme on HIV/AIDS, UNAIDS DATA 2020. 2020: Geneva.

14. Clark, R.A., et al., The impact of alternative delivery strategies for novel tuberculosis vaccines in low-income and middle-income countries: a modelling study. Lancet Glob Health, 2023. 11(4): p. e546–e555.

15. Scarponi, D., et al., Demonstrating multi-country calibration of a tuberculosis model using new history matching and emulation package - hmer. Epidemics, 2023. 43: p. 100678.

16. Iskauskas, A., hmer: History Matching and Emulation package. 2022.

17. World Health Organisation. WHO preferred product characteristics for new tuberculosis vaccines. 2018; Available from: http://apps.who.int/iris/handle/10665/273089.

18. Penn-Nicholson, A., et al., Safety and immunogenicity of candidate vaccine M72/AS01E in adolescents in a TB endemic setting. Vaccine, 2015. 33(32): p. 4025–34.

19. Kumarasamy, N., et al., Long-term safety and immunogenicity of the M72/AS01E candidate tuberculosis vaccine in HIV-positive and -negative Indian adults: Results from a phase II randomized controlled trial. Medicine (Baltimore), 2018. 97(45): p. e13120.

20. World Health Organisation, Revised BCG vaccination guidelines for infants at risk for HIV infection. Weekly Epidemiological Record, 2007. 82(21): p. 193–196.

21. UNICEF. Bacillus Calmette–Guérin (BCG) vaccine price data. 2021; Available from: https://www.unicef.org/supply/documents/bacillus-calmettegu%C3%A9rin-bcg-vaccine-price-data.

22. Gavi The Vaccine Alliance. GAVI Alliance Vaccine Introduction Grant and Operational Support for Campaigns Policy. Version 1.0. 2013; Available from: http://www.gavi.org

23. UNICEF. Costs of vaccinating a child. 2020; Available from: https://immunizationeconomics.org/recent-activity/2021/6/15/standard-costs-of-vaccinating-a-child

24. World Health, O., BCG vaccine: WHO position paper, February 2018 – Recommendations. Vaccine, 2018. 36(24): p. 3408–3410.

25. GBD 2019 Disease and Injuries Collaborators, Global burden of 369 diseases and injuries in 204 countries and territories, 1990-2019: a systematic analysis for the Global Burden of Disease Study 2019. The Lancet, 2020. 396(10258): p. 1204–1222.

26. The World Bank. World development indicators. 2022; Available from: https://data.worldbank.org/.

27. Ochalek, J., J. Lomas, and K. Claxton, Estimating health opportunity costs in low-income and middle-income countries: a novel approach and evidence from cross-country data. BMJ Glob Health, 2018. 3(6): p. e000964.

28. Portnoy, A., et al., The cost and cost-effectiveness of novel tuberculosis vaccines in low- and middle-income countries: A modeling study. PLoS Med, 2023. 20(1): p. e1004155.

29. Scarponi, D., et al., Is neglect of self-clearance biassing TB vaccine impact estimates? medRxiv, 2023.

