## Additional file 1 for "Modelling the health and economic impacts of M72/AS01_E_ vaccination and BCG-revaccination: estimates for South Africa"

Tom Sumner et al.

### SUPPLEMENTARY METHODS

### 1. Model structure

We developed an age-stratified dynamic transmission model to estimate the health impact of new of tuberculosis (TB) vaccine in South Africa.

The model is based on the model described in [1, 2] with additional structure to represent HIV infection, progression and treatment. The TB natural history structure is described below (section 1.1). The additional HIV structure is described in section 1.2. Details of the way in which vaccines are modelled are given in section 4.

#### 1.1 TB Natural history model structure

The TB structure (Figure S1.1) divides the population into eight classes (U_N_ = Uninfected-Naive; L_F_ = Latent-Fast (recently infected); L_S_ = Latent-Slow (long-term infection); L_0_ = Latent-Zero (cleared infection), D_S_ = Subclinical Disease; D_C_ = Clinical Disease; T = On-Treatment; R = Recovered). It was created by adapting features of previous models and has been described previously in [1, 2]. Individuals with no previous exposure or infection with *Mycobacterium tuberculosis* (M.tb) (U_N_) can be infected at a rate λ and progress to the Latent-Fast state (L_F_). L_F_ represents a sustained infection state (i.e. there is no direct reversion to U_N_) from which individuals may: (i) progress rapidly to Subclinical Disease (D_S_), (ii) transition to the Latent-Slow state (L_S_). Those in the L_S_ state may i) progress to D_S_ (at a lower rate than those in L_F_), be re-infected and return to L_F_ or clear their infection and transition to the L_0_ state. Individuals in the L_0_ state cannot progress directly to TB disease but can be re-infected and return to the L_F_ state. Those in the Subclinical Disease class (D_S_) are assumed to be infectious but with a reduced infectiousness compared to those with clinical disease (D_C_) and are assumed to display no symptoms of TB. These individuals may naturally cure (without treatment) to the Resolved class (R), or progress to Clinical Disease (D_C_). Those in the D_C_ state are infectious and display symptoms of tuberculosis. They may naturally cure (like those in D_S_) or be diagnosed and started on treatment (T). Those who successfully complete treatment transition to R while those who do not return to D_C_. Individuals in the R state can return to D_S_ either through relapse or reinfection. Those in the L_S_, L_0_ and R classes are partially protected against reinfection (compared to the U_N_ state). Excess mortality due to TB may occur in the D_C_, T and R states.

The model is also stratified into two groups representing different access to health care to allow us to capture the correlation between poorer access and increased TB burden. The high-access-to-care group represents the top 3 quintiles (60% of the population) and low-access-to-care group represents the bottom 2 quintiles (40% of the population). The low-access-to-care stratum has a higher force of infection per contact, a reduced rate of treatment initiation, and an increased rate of tuberculosis progression.

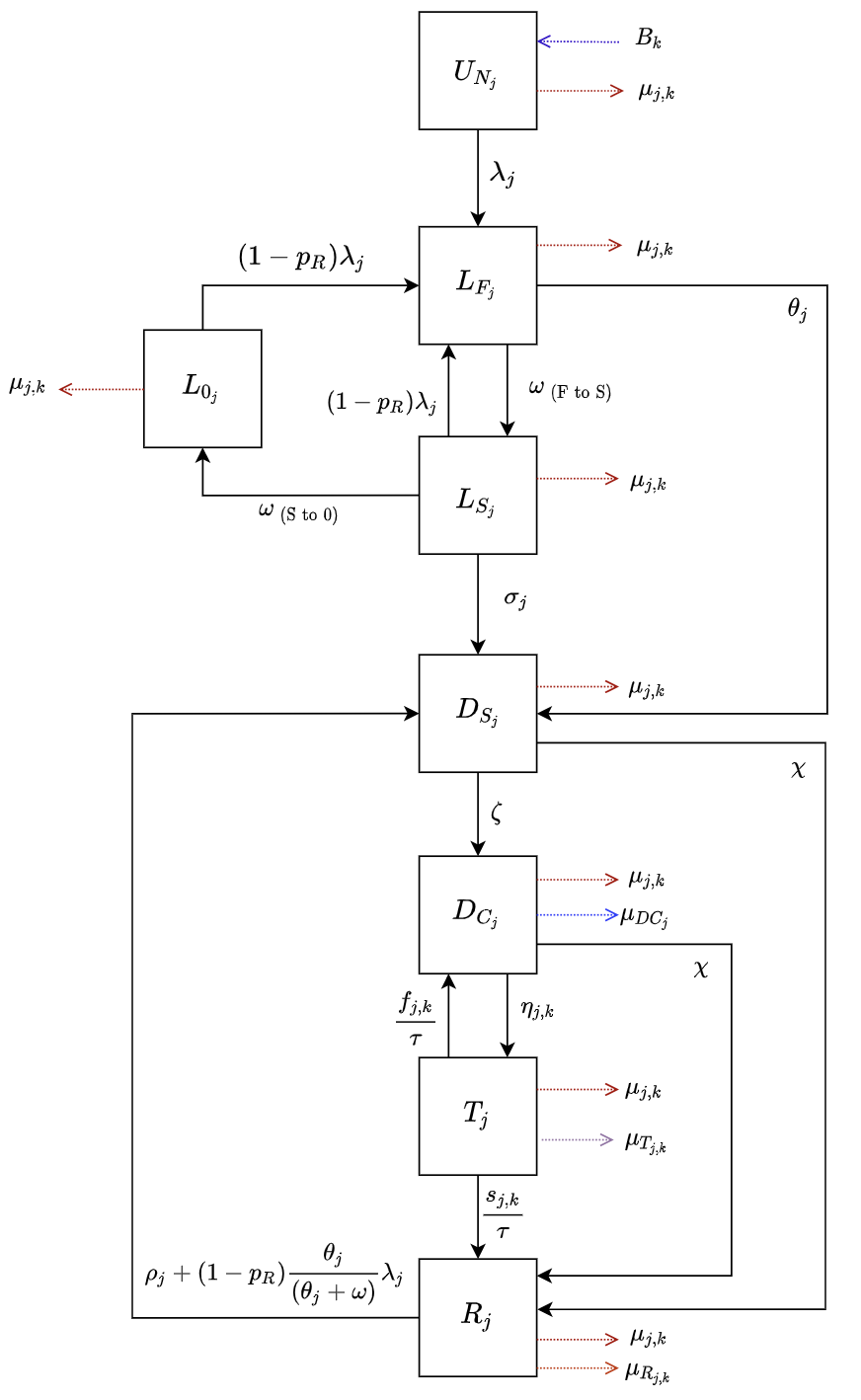

**Figure S1.1**	**Tuberculosis natural history model structure**

*Abbreviations: U_N_ = Uninfected-Naive; L_F_ = Latent-Fast; L_S_ = Latent-Slow; L_0_ = Latent-Zero, D_S_ = Subclinical Disease; D_C_ = Clinical Disease; T = On-Treatment; R = Recovered. Subscript j represents parameters that vary by age, and subscript k represents parameters that vary over time.*

#### 1.2 HIV model structure

The HIV structure (shown in Figure S1.2) is designed to capture the stages of HIV infection and treatment relevant for TB epidemiology and to allow for differences in the targeting and efficacy of TB vaccines by HIV status.

The population is divided into nine classes based on HIV status. Upon infection HIV negative individuals (H_0_) move to the undiagnosed HIV infected state with a CD4 ≥350 (H_u1_). Their CD4 can decline (to < 350) (H_u2_). HIV infected individuals can be have their HIV diagnosed, with those in H_u1_ and H_u2_, moving to the diagnosed states H_d1_ and H_d2_, respectively. Diagnosed individuals can initiate treatment and enter the ART classes (A_n1_ and A_n2_) after which they can become virally suppressed (A_s1_ and A_s2_). If ART is discontinued, individuals in the on ART-not virally suppressed classes return to the diagnosed classes. Those on ART and virally suppressed can experience a viral rebound and return to the corresponding on ART, not virally suppressed classes. There is no CD4 count decline for individuals who are virally suppressed (17). The model assumes that the rates of diagnosis, ART initiation and discontinuation and, viral suppression and rebound do not depend on CD4 count. All HIV infected individuals experience an HIV related mortality at a rate μ_i_, where the subscript i represents the relevant HIV sub-category.

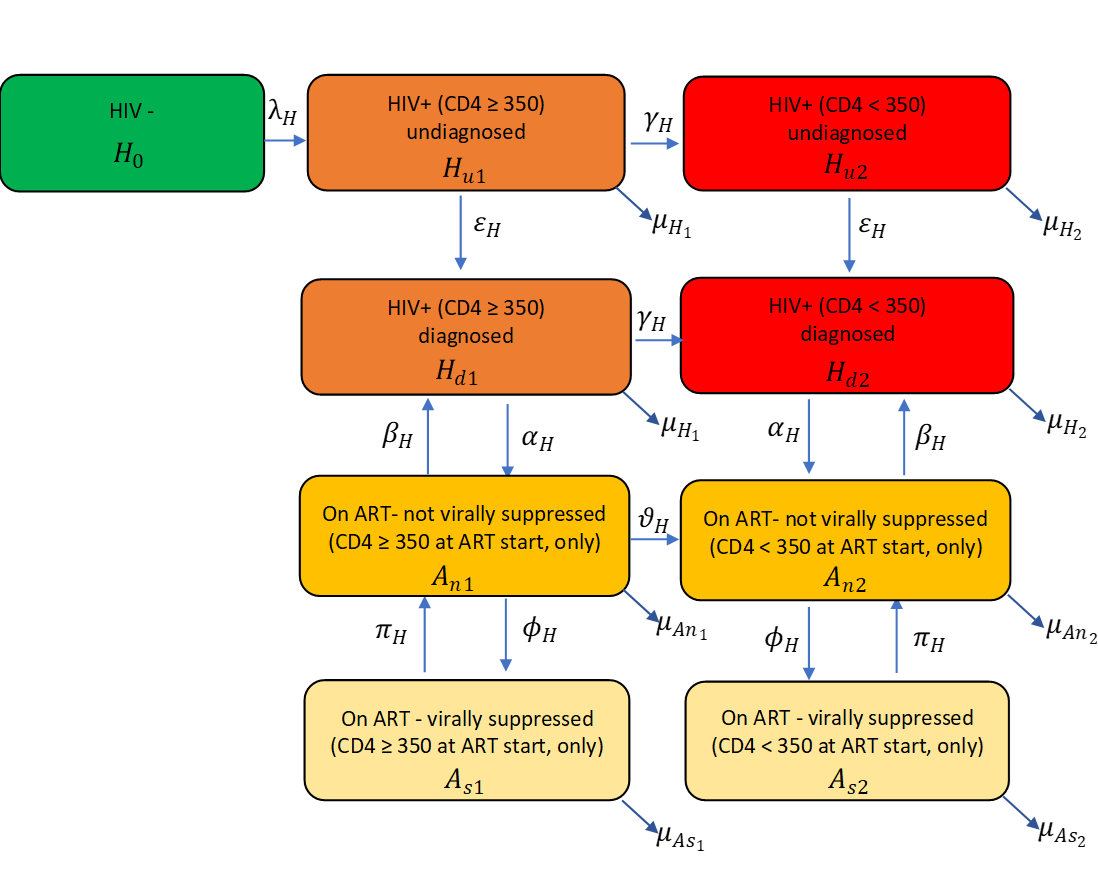

**Figure S1.2**	**HIV model structure**

### 2. Parameters

Parameters used in the TB model structure are described in section 2.1 and those used in the HIV model in section 2.2. The effects of HIV on the TB model parameters are described in section 2.3. Details on parameters related to treatment are provided in section 2.4.

These tables include definitions, sources, and information on whether the parameter is fixed or varied as well as whether they are varied by age or time during calibration.

The parameter ranges provided are priors fitted during calibration in a Bayesian analysis. We assume that all values within the prior range are equally likely. The prior ranges were pre-specified based on literature review and were reviewed as new data became available. The calibration process is described in section 3.

#### 2.1 TB model parameter values and data sources

**Table S2.1	TB model parameter values and sources**

| **Description** | **Units** | **Symbol** | **Prior** | **Fixed or Varying During Calibration** | **Age Varying** | **Time Varying** | **Source** |
| --- | --- | --- | --- | --- | --- | --- | --- |
| ***Births and deaths (excluding on-treatment mortality)*** | | | | | | | |
| Birth rate | Per year | 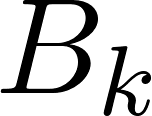<https://www.codecogs.com/eqnedit.php?latex=B_k#0> | United Nations World Population Prospects population estimates and projections | Fixed | No | Yes | [3] |
| Background mortality rate | Per year | 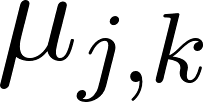<https://www.codecogs.com/eqnedit.php?latex=%5Cmu_%7Bj%2Ck%7D#0> | Calculated in the model from United Nations population estimates and projections | Fixed | Yes; age specific mortality rates from demographic dataset | Yes | [3] |
| Mortality rate for clinical tuberculosis disease | Per person  per year | 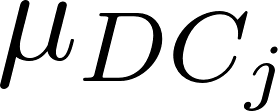<https://www.codecogs.com/eqnedit.php?latex=%5Cmu_%7BDC_j%7D#0> | (0–0.178) | Varying | Yes; value for children is greater than value for adults | No | [4] |
| Mortality rate post-tuberculosis disease | Per person  per year | 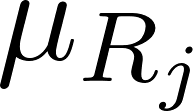<https://www.codecogs.com/eqnedit.php?latex=%5Cmu_%7BR_j%7D#0> | 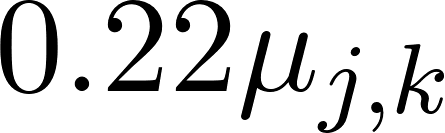<https://www.codecogs.com/eqnedit.php?latex=0.22%5Cmu_%7Bj%2Ck%7D#0> | Fixed relationship | Yes because 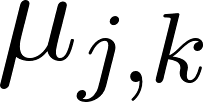<https://www.codecogs.com/eqnedit.php?latex=%5Cmu_%7Bj%2Ck%7D#0> varies | Yes because 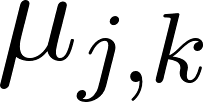<https://www.codecogs.com/eqnedit.php?latex=%5Cmu_%7Bj%2Ck%7D#0> varies | [5] |
| ***Natural History*** | | | | | | | |
| Force of infection | Per year | 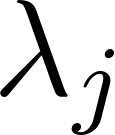<https://www.codecogs.com/eqnedit.php?latex=%5Clambda_j#0> | Calibrated in the model | Fixed Equation | Yes; age specific contact rates^23^ | No | *Calculated* |
| Probability of transmission per infectious contact | - | 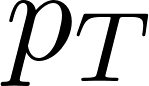<https://www.codecogs.com/eqnedit.php?latex=p_T#0> | (0–1) | Varying | No | No | *Assumed* |
| Infectiousness of subclinical relative to clinical tuberculosis | - | 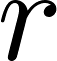<https://www.codecogs.com/eqnedit.php?latex=r#0> | 0.8 | Fixed | No | No | [6] |
| Rate of fast progression to disease, by age | Per person per year | 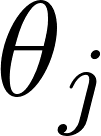<https://www.codecogs.com/eqnedit.php?latex=%5Ctheta_j#0> | (0.0696–0.111) | Varying | Yes; value for children is less than value for adults. | No | [7] |
| Rate from L_F_ to L_S_ | Per person per year | 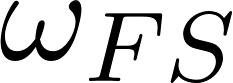<https://www.codecogs.com/eqnedit.php?latex=%5Comega_%7BFS%7D#0> | 0.5 | Fixed | No | No | *Assumed* |
| Rate of reactivation from L_S_, by age | Per person per year | 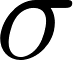<https://www.codecogs.com/eqnedit.php?latex=%5Csigma#0> | (0.000135–0.00113) | Varying | Yes; value for children is less than value for adults. | No | [7] |
| Rate from L_S_ to L_0_ | Per person per year | 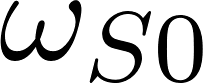<https://www.codecogs.com/eqnedit.php?latex=%5Comega_%7BS0%7D#0> | (0.0254–0.0467) | Varying | No | No | [7] |
| Rate of progression from D_S_ to D_C_ | Per person per year | 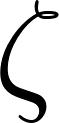<https://www.codecogs.com/eqnedit.php?latex=%20%5Czeta%20#0> | (0–12) | Varying | No | No | *Assumed* |
| Rate of natural cure from D_C_ and D_S_ | Per person per year | 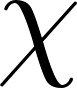<https://www.codecogs.com/eqnedit.php?latex=%5Cchi#0> | (0.10–0.25) | Varying | No | No | [8, 9] |
| Rate of relapse from R, by age | Per person per year | 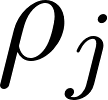<https://www.codecogs.com/eqnedit.php?latex=%5Crho_j#0> | (0.0001–0.07) | Varying | Yes; value for children is less than value for adults. | No | [10, 11] |
| ***Protection Parameters*** | | | | | | | |
| Protection from reinfection  L_S_, L_F_, L_0_, R | - | 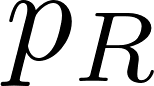<https://www.codecogs.com/eqnedit.php?latex=%20p_R%20#0> | (0.60–0.85) | Varying | No | No | [9, 12, 13] |
| Access-to-care  parameter | - | 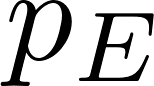<https://www.codecogs.com/eqnedit.php?latex=p_E#0> | (0–1) | Varying | No | No | *Assumed* |

Table S2.2 shows how the age varying TB parameters in table S2.1 are implemented in the model. These parameters are allowed to differ between adults (≥15 years of age) and children (<15 years of age). For the rates of reactivation, relapse, and fast progression to tuberculosis disease, we assume that the rate for children is less than that for adults. For mortality rates, we assume the rate for children is higher than that for adults.

**Table S2.2	How age varying TB parameters are operationalized**

| **Parameter** | **Prior Range** | **Age Varying Description** | **Age Scaling Parameter** | **Adults**  **(A15)** | **Children**  **(A0)** |
| --- | --- | --- | --- | --- | --- |
| 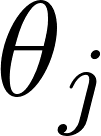<https://www.codecogs.com/eqnedit.php?latex=%5Ctheta_j#0>  Rate of fast progression to disease | 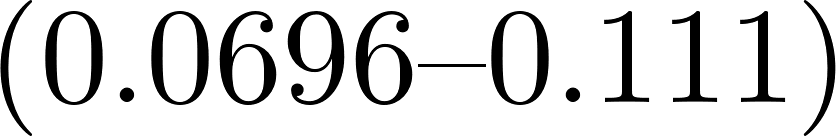<https://www.codecogs.com/eqnedit.php?latex=(0.0696%5Ctextendash0.111)#0> | Value for children is **less** than value for adults | Sample 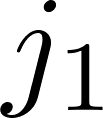  from 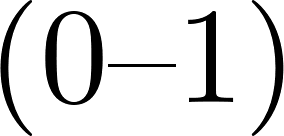<https://www.codecogs.com/eqnedit.php?latex=(0%5Ctextendash1)#0> | Sample 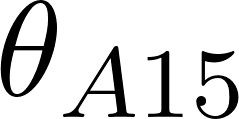<https://www.codecogs.com/eqnedit.php?latex=%5Ctheta_%7BA15%7D#0> from prior range | 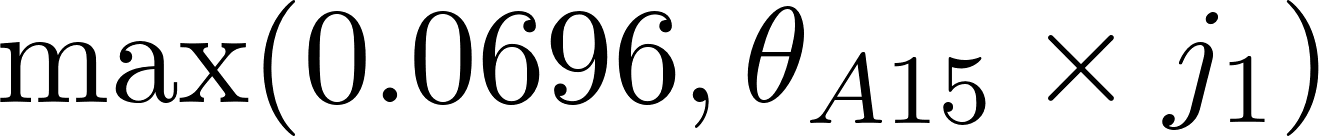<https://www.codecogs.com/eqnedit.php?latex=%5Cmax(0.0696%2C%20%5Ctheta_%7BA15%7D%20%5Ctimes%20j_1)#0> |
| 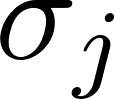<https://www.codecogs.com/eqnedit.php?latex=%5Csigma_j#0>  Rate of reactivation from L_S_ | 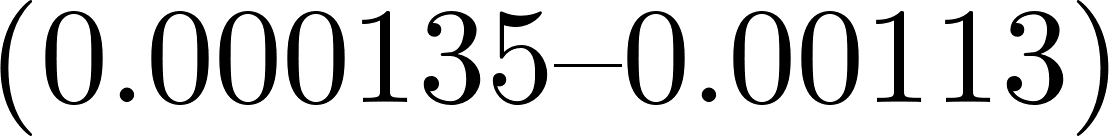<https://www.codecogs.com/eqnedit.php?latex=(0.000135%5Ctextendash0.00113)#0> | Value for children is **less** than value for adults | Sample 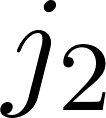<https://www.codecogs.com/eqnedit.php?latex=j_2#0>  from 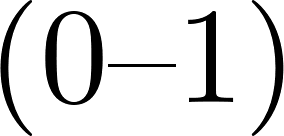<https://www.codecogs.com/eqnedit.php?latex=(0%5Ctextendash1)#0> | Sample 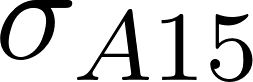<https://www.codecogs.com/eqnedit.php?latex=%5Csigma_%7BA15%7D#0> from prior range | 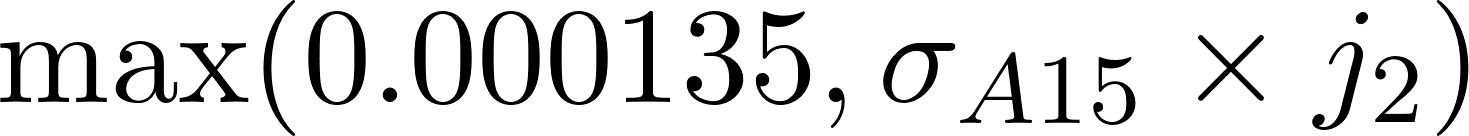<https://www.codecogs.com/eqnedit.php?latex=%5Cmax(0.000135%2C%20%5Csigma_%7BA15%7D%20%5Ctimes%20j_2)#0> |
| 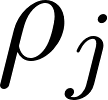<https://www.codecogs.com/eqnedit.php?latex=%5Crho_j#0>  Rate of relapse from R | 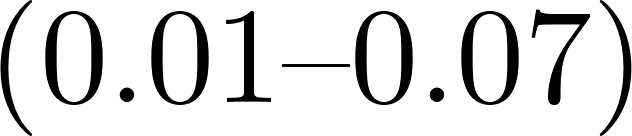<https://www.codecogs.com/eqnedit.php?latex=(0.01%5Ctextendash0.07)#0> | Value for children is **less** than value for adults | Sample 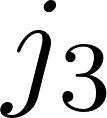<https://www.codecogs.com/eqnedit.php?latex=j_3#0>  from 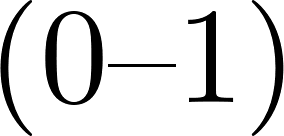<https://www.codecogs.com/eqnedit.php?latex=(0%5Ctextendash1)#0> | Sample 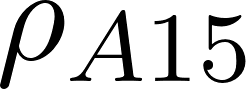<https://www.codecogs.com/eqnedit.php?latex=%5Crho_%7BA15%7D#0> from prior range | 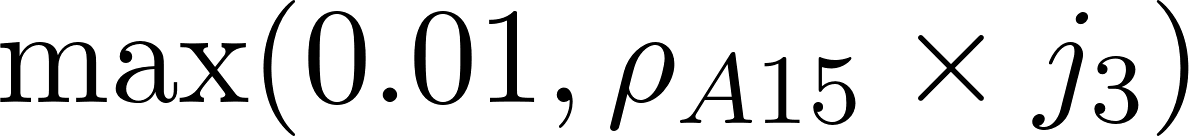<https://www.codecogs.com/eqnedit.php?latex=%5Cmax(0.01%2C%20%5Crho_%7BA15%7D%20%5Ctimes%20j_3)#0> |
| 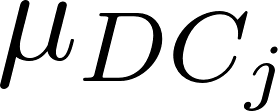<https://www.codecogs.com/eqnedit.php?latex=%5Cmu_%7BDC_j%7D#0>  Clinical TB mortality rate per year | 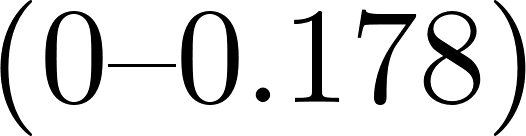<https://www.codecogs.com/eqnedit.php?latex=(0%5Ctextendash0.178)#0> | Value for children is **greater** than value for adults | Sample 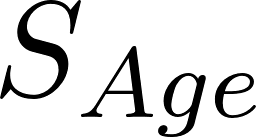<https://www.codecogs.com/eqnedit.php?latex=S_%7BAge%7D#0>  from 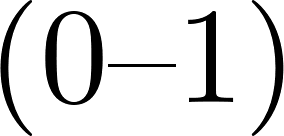<https://www.codecogs.com/eqnedit.php?latex=(0%5Ctextendash1)#0> | 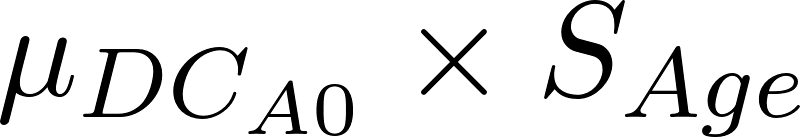<https://www.codecogs.com/eqnedit.php?latex=%5Cmu_%7BDC_%7BA0%7D%7D%20%5Ctimes%20S_%7BAge%7D#0> | Sample 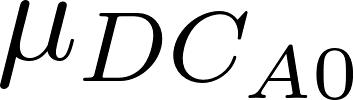<https://www.codecogs.com/eqnedit.php?latex=%5Cmu_%7BDC_%7BA0%7D%7D#0> from prior range |

#### 2.2 HIV model parameter values and data sources

**Table S2.3	HIV model parameter values and sources**

| **Description** | **Units** | **Symbol** | **Prior** | **Fixed or Varying During Calibration** | **Age Varying** | **Time Varying** | **Source** |
| --- | --- | --- | --- | --- | --- | --- | --- |
| HIV Incidence rate for uninfected individuals | Per person year | $\lambda_{H}$ | Trend data from spectrum | Varies | Yes | Yes | [14] |
| HIV progression rate from CD4 ≥ 350 to CD4 < 350 | Per year | $\gamma_{H}$ | 1/6.1 | Fixed | No | No | [15, 16] |
| HIV progression rate when on ART | Per year | $\vartheta_{H}$ | ${\gamma_{H}}/2$ | Fixed | No | No | [17, 18] |
| Rate of HIV diagnosis | Per year | $\epsilon_{H}$ | - | Varies | Yes; rate in children is 0.82 that in adults | Yes | fitted in calibration |
| Rate of treatment initiation | Per year | $\alpha_{H}$ | - | Varies | Yes; rate in children is 0.75 that in adults | Yes | fitted in calibration |
| Rate of treatment discontinuation in adults (15+) | Per year | $\beta_{H}$ | 0.074 | Fixed | Yes; different value for adults and children | No | [19, 20] |
| Rate of treatment discontinuation in children (<15) | Per year | $\beta_{H}$ | 0.092 | Fixed | Yes; different value for adults and children | No | [21-23] |
| Rate of viral suppression | Per year | $\Phi_{H}$ | - | Varies | Yes; rate in children is 0.82 that in adults | No | [14] |
| Rate of viral rebound | Per year | $\pi_{H}$ | 0.099 | Fixed | No | No | [24] |
| HIV related death rate, not on treatment  (CD4 ≥ 350) | Per year | $\mu_{H_{1}}$ | 0.0-0.012 | Varies | No | No | [25] |
| HIV related death rate, on ART, not virally suppressed  (CD4 ≥ 350) | Per year | $\mu_{{An}_{1}}$ | 0.2$\mu_{H_{1}}$ | Varies | No | No | [26] |
| HIV related death rate, on ART, virally suppressed  (CD4 ≥ 350) | Per year | $\mu_{{As}_{1}}$ | 0.32$\mu_{{An}_{1}}$ | Varies | No | No | [24] |
| HIV related death rate (CD4 < 350, not on treatment) | Per year | $\mu_{H_{2}}$ | 0-0.33 | Varies | No | No | [16] |
| HIV related death rate, on ART, not virally suppressed  (CD4 < 350) | Per year | $\mu_{{An}_{2}}$ | 0.2$\mu_{H_{2}}$ | Varies | No | No | [26] |
| HIV related death rate, on ART, not virally suppressed  (CD4 < 350) | Per year | $\mu_{{As}_{2}}$ | 0.32$\mu_{{An}_{2}}$ | Varies | No | No | [24] |

#### 2.3 Effects of HIV on TB model parameters

We assume that HIV status affects transitions in the TB model in several ways: increases in susceptibility to infection [27]; increased rates of progression to TB disease (primary, reactivation and relapse) [28, 29]; reduced rates of natural recovery; increased mortality rate associated with TB [30]. We assume that these effects depend on CD4 count, ART and viral suppression. We also assume that HIV affects the rate at which individuals with TB are diagnosed and started on treatment.

The effects of HIV on TB transitions are determined via a set of parameters that are used to scale the parameter values in the HIV uninfected strata. First the HIV uninfected values are scaled to give overall values for HIV infected populations. These scaling values are varied during calibration. Additional scaling factors are then used to adjust the values across HIV infected strata. These factors are fixed during calibration. The values are given in table S2.4.

For susceptibility to infection, progression to TB disease, TB mortality and TB treatment initiation the effects (which are to increase the rates) are implemented in the model as follows. For a given parameter x in the HIV uninfected population, the value in the specific HIV strata is given by:

$$x(1+c_{HIV}.c_{j})$$

For natural recovery from TB the effect of HIV (which is to reduce the rate) is implemented as:

$$x/(1+c_{HIV}.c_{j})$$

In both cases, $c_{HIV}$ is the overall parameter and $c_{j}$ is the strata specific scaling.

**Table S2.4	Parameters for the effect of HIV on TB transitions**

| **Overall parameters (variable)** | | |
| --- | --- | --- |
| **Transition** | **Range** | **Source** |
| Susceptibility to infection | 0-1.5 | [31] |
| Rate of progression, reactivation and relapse | 5-35 | [32-35] |
| Natural cure | 0-2 | Assumption |
| TB mortality | 1-5 | [30, 36] |
| TB treatment initiation | 0-4.5 | [37] |
| **HIV strata specific parameters (fixed)** | | |
| **HIV Strata** | **Multiplier for TB mortality** | **Multiplier for other transitions** |
| **Not on ART** | | |
| CD4 ≥ 350 | 0.8 | 1 |
| CD4 < 350 | 3.1 | 2.25 |
| **On ART, not virally suppressed** | | |
| CD4 ≥ 350 | 0.12 | 0.5 |
| CD4 < 350 | 0.4 | 0.7 |
| **On ART, virally suppressed** | | |
| CD4 ≥ 350 | 0.1 | 0.3 |
| CD4 < 350 | 0.2 | 0.4 |

#### 2.4 Treatment initiation and outcomes

The approach for simulating treatment initiation and treatment outcomes is described in detail in [1] and summarised here.

The treatment initiation rate determines the transition from clinical TB (D_c_) to the on treatment state (T). We assume that the availability of treatment started in 1960 and followed a sigmoidal increase to its current level in 2020. The rate of treatment initiation in 2020 was sampled during the calibration process. This is multiplied by the value of the sigmoidal curve (0-1) to give the value for each year. We also assume that the rate of treatment initiation is lower in children than adults. This is implemented via an additional scaling parameter sampled between 0-1 during calibration.

Our model has 3 possible outcomes while on treatment: death, treatment completion (transition to the R state), treatment non-completion (return to the D_c_ state). WHO reported data [38] on outcomes is used to derive the rates of on-treatment mortality, treatment completion and treatment-non completion. First the fraction of deaths among children (>15 years old) starting treatment is sampled during calibration. This is scaled by a multiplier to give the fraction of deaths in adults. The fraction who do not die are then divided between completion and non-completion based on the ratio of completion to completion plus non-completion reported in the WHO data. Table S2.5 below shows the calculations.

**Table S2.5**	**Calculating treatment outcome parameter values for adults and children**

| **Parameter** | **Adults (A_15_)** | **Children (A_0_)** |
| --- | --- | --- |
| 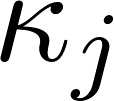<https://www.codecogs.com/eqnedit.php?latex=%5Ckappa_j#0>  On-treatment mortality fraction | 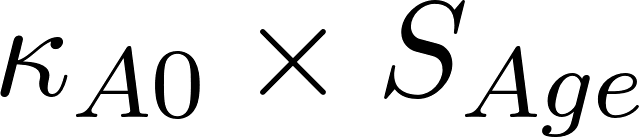<https://www.codecogs.com/eqnedit.php?latex=%5Ckappa_%7BA0%7D%20%5Ctimes%20S_%7BAge%7D#0> | Sample 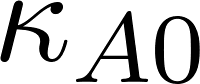<https://www.codecogs.com/eqnedit.php?latex=%5Ckappa_%7BA0%7D#0> from (0-0.437) |
| 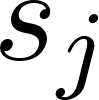<https://www.codecogs.com/eqnedit.php?latex=s_j#0>  On-treatment completion fraction | 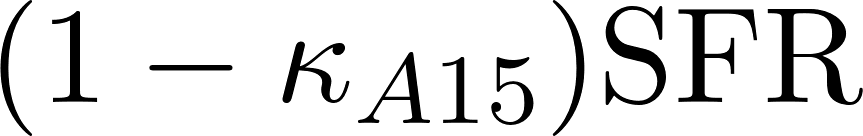<https://www.codecogs.com/eqnedit.php?latex=(1-%5Ckappa_%7BA15%7D)%5Ctext%7BSFR%7D#0> | 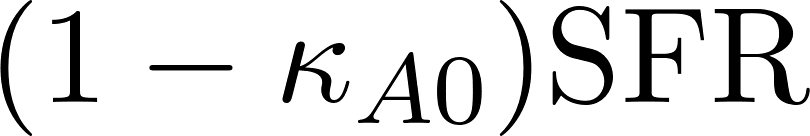<https://www.codecogs.com/eqnedit.php?latex=(1-%5Ckappa_%7BA0%7D)%5Ctext%7BSFR%7D#0> |
| 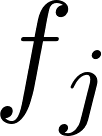<https://www.codecogs.com/eqnedit.php?latex=f_j#0>  On-treatment non-completion fraction | 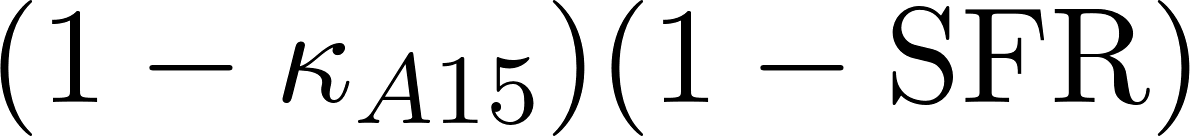<https://www.codecogs.com/eqnedit.php?latex=(1-%5Ckappa_%7BA15%7D)(1-%5Ctext%7BSFR%7D)#0> | 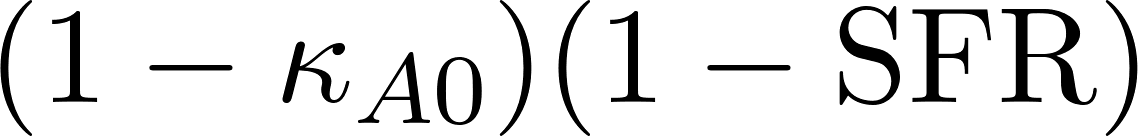<https://www.codecogs.com/eqnedit.php?latex=(1-%5Ckappa_%7BA0%7D)(1-%5Ctext%7BSFR%7D)#0> |

### 3. Model simulation and calibration

#### 3.1 Model simulation

The model consists of a system of ordinary differential equations which define the derivatives with respect to time of the state variables. The transitions between state variables, together with the parameters and other model inputs, are specified in an xml file using a standardised schema. The equations are then generated and solved using a matrix multiplication approach. This is implemented in the R programming language [39].

The model is initialised by distributing the population between the TB disease states using a parameter representing the proportion of the population uninfected at the start of the simulation. Forty percent of the population was assigned to the low access-to-care stratum and the remaining sixty percent of the population was assigned to the high access-to-care stratum.

The model is used to simulate the progression of the TB and HIV epidemics from 1900 to 2050. The model is initially stimulated without new TB vaccines to establish the baseline no-new-vaccine scenario. For each year of the baseline simulation the birth rate and age-sepcific mortality rates are adjusted to match the UN population estimates and projections. This allows us to account for TB and HIV mortality, which is simulated independently in our model but implicity included in the UN estimates and projections. The baseline, with the adjusted birth and mortality rates, is then used to simulate different vaccination scenarios.

#### 3.2 Model calibration

To establish the no-new-vaccine baseline the model was calibrated to a set of TB and HIV calibration targets (see table S3.1). Calibration was carried out using history matching with emulation, a method that allows us to explore high-dimensional parameter spaces efficiently and robustly [40, 41]. History matching progresses as a series of iterations, called waves, where implausible areas of the parameter space, i.e., areas that are unable to give a match between the model output (e.g., the predicted incidence rate by the model) and the empirical data (e.g., the incidence rate calibration target from the WHO data), are found and discarded. In order to identify implausible parameter sets, emulators, which are statistical approximations of model outputs that are built using a modest number of model runs, are used. Emulators provide an estimate of the value of the model at any parameter set of interest, with the advantage that they are orders of magnitude faster than the model. History matching with emulation, implemented through the *hmer* package in R [42], considerably reduced the size of the parameter space to investigate. Rejection sampling was then performed on the reduced space to identify at least 1000 parameter sets that matched all targets. Once we had obtained 1000 parameter sets that produced output consistent with the calibration targets, we used those parameter sets with the mechanistic model to simulate the future trajectories of TB. Our future projection assumed that TB and HIV diagnosis and treatment continued at 2020 levels, and that HIV incidence remained constant at 2020 estimates.

**Table S3.1	South Africa model calibration targets**

| **Calibration Targets** | **Year** | **HIV status** | **Age (years)** | **Estimate (lower-upper bound)** | **Source** |
| --- | --- | --- | --- | --- | --- |
| **Tuberculosis** | | | | | |
| TB incidence rate (per 100,000) | 2000 | All | All | 762 (475-1120) | WHO TB report 2021  [43] |
|  |  | HIV+ |  | 559 (318-866) |  |
|  | 2020 | All | All | 615 (427-835) |  |
|  |  |  | 0-14 | 224 (142-306) |  |
|  |  |  | 15+ | 774 (488-1060) |  |
|  |  | HIV+ | All | 357 (28-486) |  |
| TB mortality rate (per 100,000) | 2000 | All | All | 317 (199-461) |  |
|  |  | HIV+ |  | 258 (144-405) |  |
|  | 2020 | All |  | 99 (59-150) |  |
|  |  | HIV+ |  | 62 (25-115) |  |
| TB notification rate (per 100,000)* | 2000 | All | All | 336 (269-403) |  |
|  | 2020 |  | 0-14 | 97 (78-116) |  |
|  |  |  | 15+ | 464 (371-557) |  |
| TB prevalence (per 100,000) | 2019 | All | All | 737 (580-890) | South Africa TB prevalence survey  [44] |
| Percentage of TB prevalence that is subclinical (%) | 2019 | All | All | 57.7 (51.1-64.1) |  |
| TB prevalence ratio (high to low access to care) | 2019 | All | All | 0.691 (0.591-0.791) | [45, 46] |
| **HIV** | | | | | |
| HIV prevalence (%) | 2000 | - | 0-14 | 0.9 (0.5-1.2) | UNAIDS Data 2021  [47] |
|  |  |  | 15+ | 10.1 (6.1-13.8 |  |
|  | 2020 |  | 0-14 | 1.8 (1.2-3.2) |  |
|  |  |  | 15+ | 17.7 (11.7-22.5) |  |
| Percentage of PLHIV who know their status (%) | 2020 |  | 0-14 | 75 (47-100) |  |
|  |  |  | 15+ | 93 (62-100) |  |
| Percentage of PLHIV who are on ART (%) | 2010 |  | 0-14 | 24 (14-34) |  |
|  |  |  | 15+ | 23 (15-31) |  |
|  | 2020 |  | 0-14 | 47 (30-81) |  |
|  |  |  | 15+ | 73 (48-93) |  |
| Percentage of people on ART who are virally suppressed | 2020 |  | 0-14 | 70 (44-100) |  |
|  |  |  | 15+ | 92 (61-100) |  |
| HIV mortality rate (per 100,000) | 2000 |  | All | 289 (162-445) |  |
|  | 2020 |  |  | 140 (73-253) |  |

*Notifications are reported as an absolute number of cases (point estimate without ranges). Notification rates were calculated using UN Population estimates as denominator. The range was calculated assuming +/- 20% of the reported notifications to account for possible underreporting of treatment initiation or inclusion of false positive cases.

### 4. Vaccines

#### 4.1 Classification of tuberculosis vaccines

Following our previous modelling of novel TB vaccines [1, 2] we characterised vaccines on four key characteristics: the vaccine efficacy, the duration of protection, the host infection status required for the vaccine to be efficacious and the mechanism of effect.

Vaccine efficacy was modelled as “degree” where the vaccine offers partial protection to all individuals who received the vaccine. The duration of protection represents the length of time following vaccination that individuals are protected.

The host infection status required for the vaccine to be efficacious defines the *M.tb* infection status required at the time an individual is vaccinated for the vaccine to be efficacious. We define 3 types of vaccine:

i) No Current Infection (NCI) - the vaccine is efficacious in uninfected populations only
ii) Current Infection (CI) - the vaccine is efficacious in populations with current infection with *M.tb* only
iii) Any Infection (AI) - the vaccine is efficacious in both uninfected and currently infected populations

The mechanism of effect determines the type of protection provided by the vaccine. We define 3 types of vaccine:

i) Prevention of infection (POI) vaccine – protects individuals from initial or re-infection with *M.tb*
ii) Prevention of disease (POD) vaccine – protects individuals from progressing to active disease if currently or subsequently infected with M.tb.
iii) Prevention of infection and disease vaccine (POID) prevents both infection and disease.

Full details of how this different vaccine characteristics are implemented in the model can be found in [1]. We made the following additional assumptions about vaccination:

- There is no pre-vaccination infection testing so both uninfected and infected individuals will receive all vaccine types.
- Vaccines that protect against infection work by reducing the rate of both first and repeat infections
- Vaccines that protect against progression to disease work by reducing the rate of progression to subclinical disease.
- For vaccines that protect against both infection and disease we assume that the efficacy is the same for both mechanisms.

#### 4.2 M72/AS01_E_ and BCG-revaccination scenarios

For each of the vaccine product considered in the analysis we defined a “Basecase” scenario which species the 4 key vaccine characteristics described above together with the implementation year and vaccine coverage. These were based on clinical trial data [48, 49], WHO preferred product characteristics for new TB vaccines [50] and expert opinion.

We then defined a discrete set of vaccine policy options that could be considered by decision makers. These represented different choices of age targeting and inclusion of PLHIV. Finally, we defined a series of alternative values and assumption for the vaccine characteristics, implementation year and coverage. These were varied in a univariate analysis. Details of these assumptions are given in table S4.1

The Basecase introduction years (2025 and 2030 for BCG-revaccination and M72/AS01_E_ respectively) were based on considering when new trial data would become available, as well as incorporating time for licensure and policy change. The introduction year considered in scenario analyses, 2029 and 2034 for BCG-revaccination and M72/AS01_E_ respectively, was based on expert advice to the earliest possible introduction year.

The Basecase age was informed by ages of trial participants and expert advice. Additional scenarios were informed by work conducted by Pelzer et. al [51] and expert advice.

**Table S4.1	M72/AS01_E_ and BCG-revaccination scenarios evaluated in the analysis**

| **Characteristic** | **M72/AS01_E_ vaccines** | | **BCG-revaccination vaccines** | |
| --- | --- | --- | --- | --- |
|  | **Basecase** | **Univariate scenario analyses** | **Basecase** | **Univariate scenario analyses** |
| **Policy Options** | | | | |
| Age targeting | Campaign for ages 16-34, routine age 15 | Campaign for ages 18-55  Campaign for ages 11-34, routine age 10 | Campaign for ages 11-18, routine age 10 | Campaign for ages 16-34, routine age 15 |
| HIV targeting | All | All | None | On ART, ages 11-18  On ART, ages 10+  VS, ages 10+ |
| **Vaccine Characteristics and Coverage** | | | | |
| Efficacy in HIV uninfected | 50% | 60%  70% | 45% | 70% |
| Relative efficacy in PLHIV | *Medium:*  VS, 90%  NVS, 54% | *Low:*  VS, 80%  NVS, 16%  *High:*  VS, 100%  NVS, 100% | - | 100% |
| Duration of protection | 10 years | 5 years  15 years  20 years | 10 years | 5 years  15 years  20 years |
| Mechanism of effect | Prevents disease | Prevents infection and disease | Prevents infection | Prevents infection and disease |
| Host infection status | Any infection (current / no current infection) | Current infection only | No current infection only | Any infection (current / no current infection) |
| Introduction year | 2030 | 2034 | 2025 | 2029 |
| Coverage | *Medium:*  80% routine  70% campaign | *Low:*  70% routine  50% campaign  *High:*  90% routine  90% campaign | *Medium:*  80% routine  80% campaign | *Low:*  70% routine  70% campaign  *High:*  90% routine  90% campaign |

#### 4.3 Efficacy of vaccines in PLHIV

The efficacy of novel vaccines in PLHIV is uncertain. We therefore considered different assumptions about the targeting and efficacy of vaccines by HIV status. Based on expert opinion we considered 3 populations in which efficacy could differ: HIV uninfected; HIV infected and virally suppressed; all other HIV infected individuals (i.e. not virally suppressed). We assumed that the effect of HIV infection was to reduce the efficacy of vaccines and that there would be no effect on the duration of protection. We also assumed that the effect on efficacy would be the same for protection against infection or disease.

Our base case M72/AS01_E_ vaccination scenario assumed a reduced efficacy in PLHIV. We also simulated scenarios with 1) the same efficacy in PLHIV and HIV uninfected individuals (optimistic case) 2) lower efficacy in PLHIV (pessimistic case). As there is no primary data on the efficacy of M72/AS01_E_ vaccination on the incidence of TB disease in PLHIV we used data on immunogenicity of M72/AS01_E_ in PLHIV [52] to provide plausible ranges for the efficacy of vaccination. Table S4.2 gives values for the relative efficacy of M72/AS01_E_ in PLHIV (compared to HIV uninfected individuals) and details of their derivation.

**Table S4.2 Effects of HIV on efficacy of M72/AS01_E_**

| **Population** | **Base case** | **Optimistic case** | **Pessimistic case** |
| --- | --- | --- | --- |
| Virally suppressed vs HIV- | 90%  Geometric mean concentration of M72-specific IgG antibodies at 3yrs post vaccination. HIV- = 24.3  HIV+; ART+ = 22 | 100%  Seropositivity rates for M72-specific IgG antibodies at 3yrs post vaccination.  HIV- = 97.3  HIV+; ART+ = 97.1 | 80%  Geometric mean concentration of M72-specific IgG antibodies at 2yrs post vaccination. HIV- = 23.9  HIV+; ART+ = 19.1 |
| Not virally suppressed vs virally suppressed | 60%  Seropositivity rates for M72-specific IgG antibodies at 3yrs post vaccination.  HIV+; ART+ = 97.1  HIV+; ART- = 67.1 | 100%  Assumption | 16%  Geometric mean concentration of M72-specific IgG antibodies at 3yrs post vaccination. HIV+; ART+ = 22  HIV+; ART- = 22 |
| Not virally suppressed vs HIV- | 54%  Product of values above | 100%  Product of values above | 16%  Product of values above |

For BCG-revaccination our primary assumption was that BCG is only given to HIV uninfected individuals due to the risks associated with giving BCG to this population [53]. There is evidence that the benefit of BCG vaccination in individuals established on ART may outweigh the risks [54] so we explored additional scenarios in which BCG revaccination was extended to this population. In all cases we assumed the efficacy of BCG in PLHIV was the same as in HIV uninfected individuals. Table S4.3 lists the scenarios.

**Table S4.3 Scenarios for BCG-revaccination in PLHIV**

| **HIV status** | **Age range** | **Efficacy (relative to HIV-)** |
| --- | --- | --- |
| On ART | 11-18 years old | 100% |
| On ART | >10 years old | 100% |
| Virally suppressed | >10 years old | 100% |

### 5. Economic analysis methods

This section provides details of the economic analysis methods.

#### 5.1 Calculation of disability-adjusted life years

We calculated the disability-adjusted life years (DALYs) averted by vaccination from vaccine introduction to 2050 for each scenario compared to the no-new-vaccine baseline. We took disability weights for tuberculosis disease from the Global Burden of Disease 2019 study [55] and country- and age-specific life expectancy estimates from the United Nations Development Programme [3]. To incorporate parameter uncertainty in years lost due to disability (YLD) weight estimates, we made 1000 draws from disability weight uncertainty ranges.

#### 5.2 Tuberculosis-related cost model

Health system unit costs (TB diagnosis and treatment and ART), patient costs and productivity losses were identified from published literature. TB costs were estimated separately for drug susceptible and drug resistant cases. We assumed that uncertainty in costs were gamma distributed around plausible unit cost estimates – these were incorporated in the analysis in a probabilistic sensitivity analysis.

#### 5.3 Vaccine cost model

Based on expert opinion from funders, for the M72/AS01_E_ vaccine we assume a $2.50 per-dose vaccination price with two doses per course assumed in the Basecase. Based on the average estimated BCG price from 2020–2023 from UNICEF [56], the vaccine price per dose for BCG-revaccination was set at $0.17, with one dose assumed per course.

There is uncertainty in the likely costs of supply and introduction of novel TB vaccines and the costs of mass vaccination in adolescents and adult population as simulated here. We used data on delivery of other vaccines to inform our assumptions. As for the TB costs, uncertainty in vaccine cost estimates is characterised through gamma distributions.

One-time vaccine introduction costs are included in years where there is a campaign and represent non-recurring costs such as establishing infrastructure and providing training for healthcare professionals. The costs were assumed to be $2.40 (1.20–4.80) per individual in the targeted age group (as opposed to the actual number of recipients) based on the vaccine introduction support policy of Gavi, the Vaccine Alliance [57]. Vaccine delivery was assumed to be $2.50 (1.00–5.00) per dose, with a further $0.11 (0.06–0.22) supply costs per dose [58]. The cost of recipient vaccination time was $0.68 (0.29–4.00), which was calculated by multiplying a wage proxy of GDP per capita for South Africa by an estimate of the time required for vaccination [59, 60]. We assume a 5% wastage rate.

#### 5.4 Cost-effectiveness analysis and willingness-to-pay thresholds

We calculated the incremental cost effectiveness ratio as the ratio between the incremental benefit, in DALYs averted, and the incremental cost, in USD, for each run across vaccination and baseline scenario. Both costs and benefits were discounted to 2025 (when vaccination began) at 3% per year, per guidelines. We measured cost-effectiveness by 2050 against three South Africa specific cost thresholds: 1x gross domestic product (GDP) per-capita (US$5742) [60], and two country-level opportunity cost thresholds defined by Ochalek et al [the upper (US$3334), and lower (US$2480) bounds] [61].

#### 5.5 Total costs from the health-system and societal perspectives

The following costs are included in the health-system perspective:

- Vaccine costs: One-time vaccine introduction costs, recurring vaccine delivery costs, vaccine price per dose, and supply costs
- Cost of testing and diagnosis for drug-susceptible and drug-resistant cases
- Cost of treatment for drug-susceptible and drug-resistant cases

In addition to the costs from the health-system perspective, costs from the societal perspective include:

- Vaccine costs: Patient time cost for vaccination
- Non-medical patient costs (including transportation) for drug-susceptible and drug-resistant cases
- Indirect patient costs for drug-susceptible and drug-resistant cases

**Table S5.1**	**Tuberculosis testing, diagnostic, and vaccination related cost inputs**

| **Unit Cost** | **Estimate** | **Lower Bound** | **Upper Bound** | **Sources** |
| --- | --- | --- | --- | --- |
| Unit cost of testing/diagnosis for DS cases per person | $5.72 | $4.32 | $15.66 | [62] |
| Unit cost of testing/diagnosis for DR cases per person | $19.85 | $18.64 | $21.85 |  |
| Unit cost of treatment for DS cases per person | $188.70 | $106.36 | $328.22 | [63] |
| Unit cost of treatment for DR cases per person | $5701.82 | $4561.45 | $6842.18 | [64, 65] |
| Non-medical patient cost per DS-TB disease episode (including transportation) per person | $160.44 | $128.35 | $162.16 | [66] |
| Indirect patient cost per DS-TB disease episode (time spent on treatment and transport * wage) per person | $304.44 | $243.55 | $306.16 |  |
| Non-medical patient cost per DR-TB disease episode (including transportation) per person | $449.24 | $359.39 | $454.06 |  |
| Indirect patient cost per DR TB disease episode (time spent on treatment and transport * wage) per person | $852.44 | $681.95 | $857.26 |  |
| Annual ART cost per patient | $274.14 | $219.32 | $328.97 | [67] |
| Recurrent vaccine delivery cost per person per dose | $2.50 | $1.00 | $5.00 | [58] |
| One-time vaccine introduction costs per targeted person | $2.40 | $1.20 | $4.80 | [57] |
| Vaccine supply costs per person per dose | $0.11 | $0.06 | $0.22 | [58] |
| Cost of vaccination time per person per dose | $0.68 | $0.29 | $4.00 | [59, 60] |

### 6. Health impact outcomes

The following measures were calculated for each vaccine scenario as the median and 95% uncertainty range:

-	Percent incidence rate reduction in 2050 for each vaccine scenario compared to the estimated value in 2050 by *No-New-Vaccine* baseline

-	Percent mortality rate reduction in 2050 for each vaccine scenario compared to the estimated value in 2050 by *No-New-Vaccine* baseline

-	Cumulative cases averted for each vaccine scenario between vaccine introduction (either 2025 or 2030) and 2050 compared to the cumulative number of cases estimated by the *No-New-Vaccine* baseline between the corresponding years

-	Cumulative deaths averted for each vaccine scenario between vaccine introduction (either 2025 or 2030) and 2050 compared to the cumulative number of cases estimated by the *No-New-Vaccine* baseline between the corresponding years

-	Cumulative treatments averted for each vaccine scenario between vaccine introduction (either 2025 or 2030) and 2050 compared to the cumulative number of cases estimated by the *No-New-Vaccine* baseline between the corresponding years

### SUPPLEMENTARY RESULTS

### 7. No-new-vaccine baselines

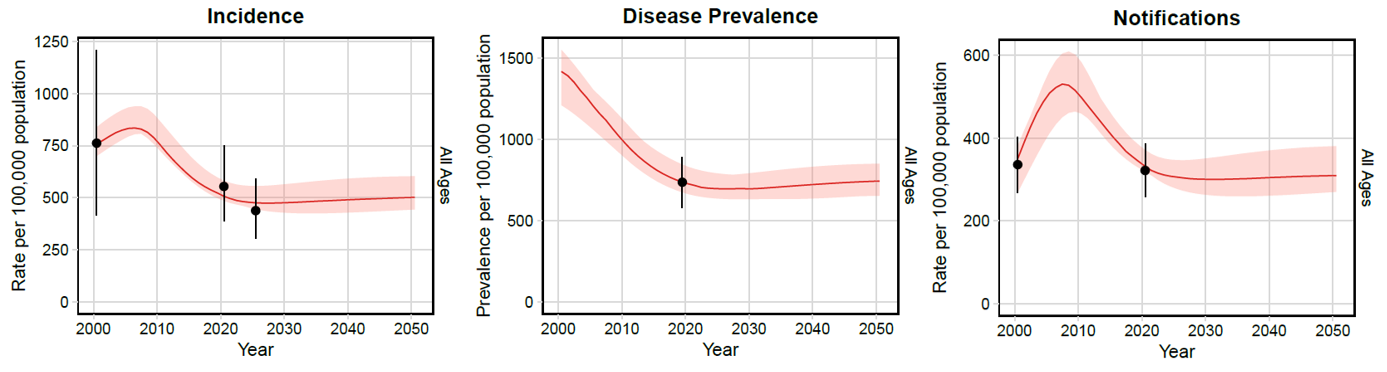

**Figure S7.1**	**Tuberculosis incidence, disease prevalence and case notification rate trends from 2000–2050 for all ages**

*The red trend line indicates the median modelled output with 95% uncertainty shown by the shaded area. The dot and vertical line is the calibration target from Table S3.1.*

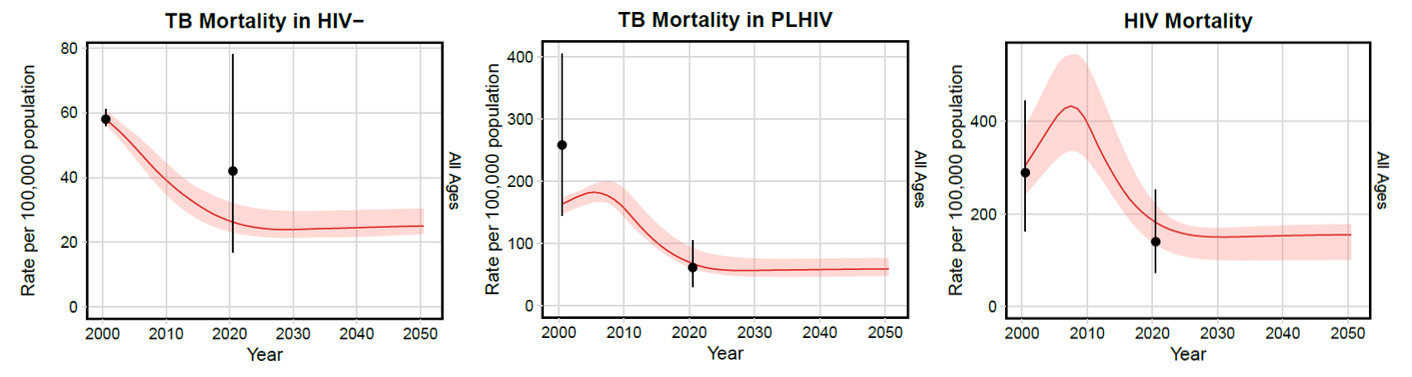

**Figure S7.2**	**Tuberculosis mortality (by HIV status) and HIV mortality rate trends from 2000–2050 for all ages**

*The red trend line indicates the median modelled output with 95% uncertainty shown by the shaded area. The dot and vertical line is the calibration target from Table S3.1.*

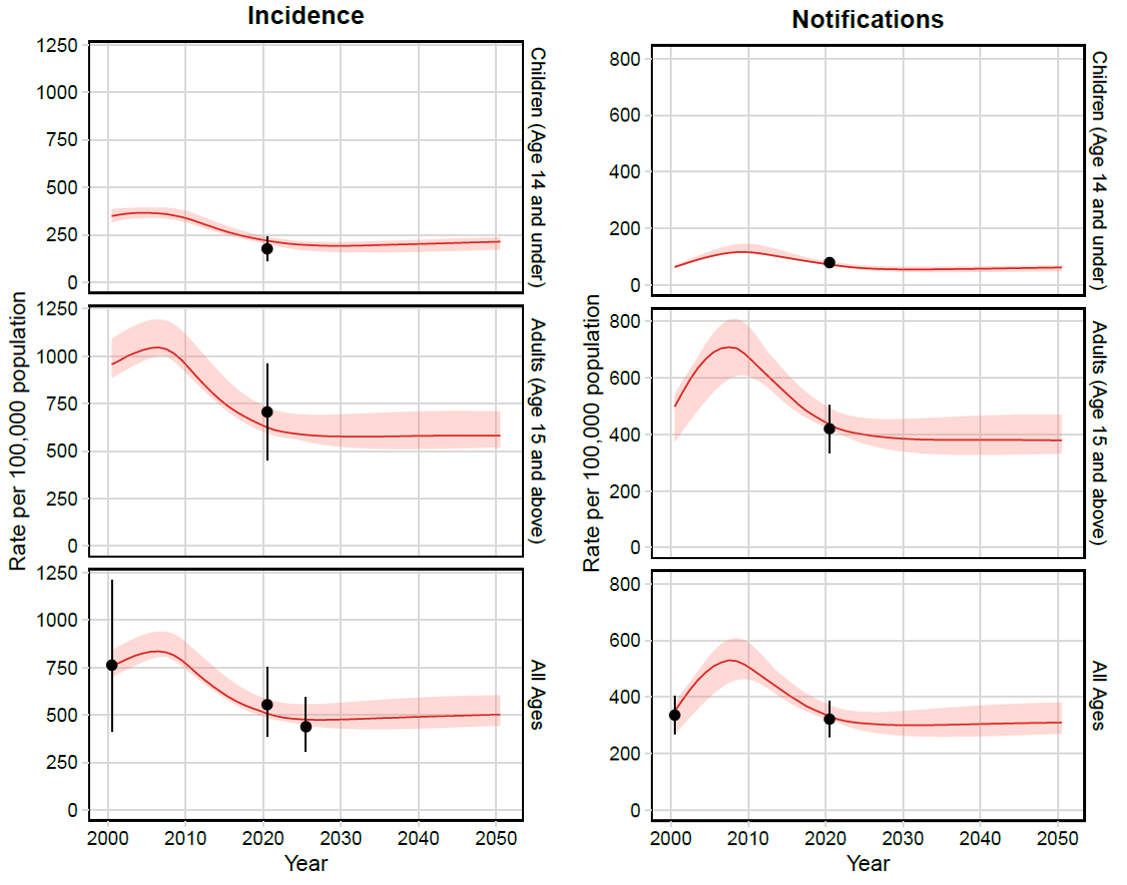

**Figure S7.2**	**Tuberculosis incidence and notification rate trends from 2000–2050 by age**

*The red trend line indicates the median modelled output with 95% uncertainty shown by the shaded area. The dot and vertical line is the calibration target from Table S3.1.*

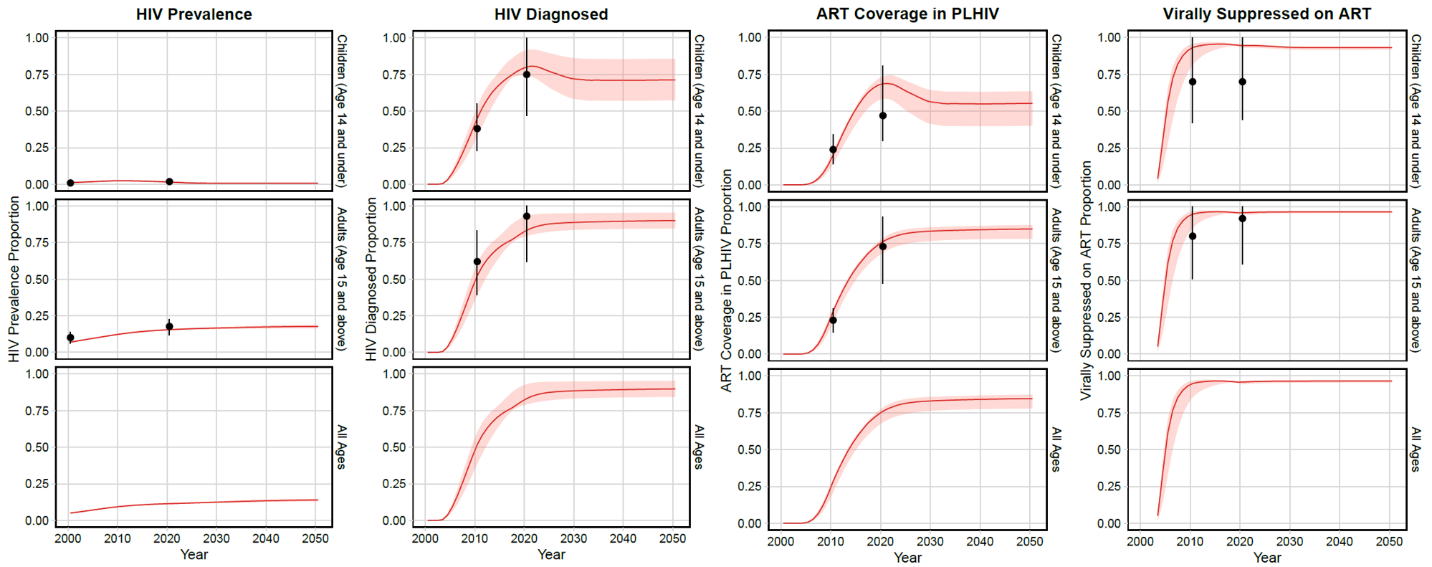

**Figure S7.2**	**HIV prevalence, percent diagnosed, ART coverage and viral suppression from 2000–2050 by age**

*The red trend line indicates the median modelled output with 95% uncertainty shown by the shaded area. The dot and vertical line is the calibration target from Table S3.1.*

### 8. Health impact results

**Table S8.1** Health impact results for the M72/AS01_E_ scenarios

| **Scenario** | **IRR in 2050**  **(%)** | **MRR in 2050**  **(%)** | **Cumulative  cases  averted**  **(millions)**  **2030–2050** | **Cumulative treatments averted**  **(millions)**  **2030–2050** | **Cumulative deaths  averted**  **(millions)**  **2030–2050** |
| --- | --- | --- | --- | --- | --- |
| Basecase | 30.1  (27.7, 34.1) | 28.3% (26.1, 32.2) | 1.56  (1.44, 1.87) | 0.84  (0.71, 0.97) | 0.22  (0.18, 0.25) |
| **Policy Scenarios** | | | | | |
| Younger ages (routine age 10, campaign for ages 11-34) | 33.2  (31.0, 37.2) | 31.6% (29.7, 35.2) | 1.72  (1.60, 2.03) | 0.89  (0.78, 1.04) | 0.24  (0.21, 0.27) |
| Older ages (campaign for ages 18-55) | 23.2  (20.9, 25.8) | 24.6% (22.1, 26.8) | 1.59  (1.49, 1.85) | 0.93  (0.82, 1.07) | 0.24  (0.2, 0.27) |
| **Vaccine Characteristic and Coverage Scenarios** | | | | | |
| 60% efficacy | 35.1  (32.4, 39.5) | 33.1  (30.6, 37.5) | 1.83  (1.69, 2.19) | 0.98  (0.84, 1.14) | 0.26  (0.21, 0.29) |
| 70% efficacy | 39.7  (36.8, 44.5) | 37.5  (34.8, 42.3) | 2.09  (1.92, 2.50) | 1.12  (0.96, 1.31) | 0.29  (0.24, 0.33) |
| Lower efficacy in PLHIV | 28.9  (26.5, 32.7) | 26.9  (24.6, 30.7) | 1.49  (1.37, 1.78) | 0.78  (0.67, 0.91) | 0.2  (0.17, 0.23) |
| Same efficacy in PLHIV | 31.8  (29.5, 36.2) | 30.5  (28.3, 34.7) | 1.68  (1.54, 1.99) | 0.92  (0.78, 1.06) | 0.24  (0.2, 0.28) |
| 5 years protection | 19.5  (17.6, 22.3) | 18.9  (17.2, 21.7) | 1.15  (1.06, 1.38) | 0.62  (0.53, 0.72) | 0.16  (0.13, 0.19) |
| 15 years protection | 35.3  (32.7, 39.8) | 32.9  (30.5, 37.2) | 1.75  (1.61, 2.09) | 0.93  (0.8, 1.09) | 0.24  (0.2, 0.28) |
| 20 years protection | 38.4  (35.8, 43.2) | 35.5  (33.1, 40.1) | 1.86  (1.71, 2.22) | 0.99  (0.85, 1.15) | 0.26  (0.22, 0.29) |
| Prevention of infection and disease | 41.4  (38.4, 47) | 39.4  (36.6, 44.7) | 2.14  (1.97, 2.55) | 1.17  (0.99, 1.35) | 0.31  (0.25, 0.35) |
| Efficacious with current infection at vaccination | 16  (14.9, 18.1) | 15.7  (14.7, 17.6) | 0.94  (0.86, 1.14) | 0.51  (0.44, 0.6) | 0.13  (0.11, 0.16) |
| 2034 introduction | 29.8  (27.6, 33.9) | 27  (25, 30.8) | 1.15  (1.05, 1.38) | 0.58  (0.49, 0.68) | 0.15  (0.12, 0.17) |
| Lower coverage | 24.5  (22.4, 27.9) | 22.9  (21, 26.2) | 1.25  (1.15, 1.50) | 0.67  (0.57, 0.77) | 0.17  (0.14, 0.2) |
| Higher coverage | 35.3  (32.7, 39.8) | 33.4  (30.9, 37.9) | 1.86  (1.71, 2.22) | 1.00  (0.85, 1.16) | 0.26  (0.22, 0.3) |

*Abbreviations: IRR = incidence rate reduction, MRR = mortality rate reduction.*

**Table S8.2** Health impact results for the BCG-revaccination scenarios

| **Scenario** | **IRR in 2050  (%)** | **MRR in 2050  (%)** | **Cumulative  cases averted**  **(millions)  2025–2050** | **Cumulative treatments averted**  **(millions) 2025–2050** | **Cumulative deaths averted**  **(millions)**  **2025–2050** |
| --- | --- | --- | --- | --- | --- |
| Basecase | 13.7  (11.8, 16.3) | 12.5  (10.8, 15) | 0.86  (0.80, 0.97) | 0.45  (0.38, 0.5) | 0.12  (0.1, 0.13) |
| **Policy Scenarios** | | | | | |
| Older ages (routine age 15, campaign for ages 16-34) | 14  (12.1, 18.5) | 12.9  (11.3, 17.1) | 0.86  (0.77, 1.02) | 0.47  (0.38, 0.54) | 0.13  (0.1, 0.14) |
| Basecase plus on ART ages 11-18 | 14.1  (12.1, 16.6) | 12.9  (11.1, 15.3) | 0.88  (0.82, 1.00) | 0.46  (0.39, 0.52) | 0.12  (0.11, 0.14) |
| Basecase plus on ART ages 10+ | 15.7  (13.5, 18.3) | 14.6  (12.5, 17.1) | 0.99  (0.92, 1.13) | 0.52  (0.46, 0.60) | 0.14  (0.12, 0.16) |
| Basecase plus virally suppressed ages 10+ | 15.6  (13.4, 18.2) | 14.5  (12.4, 17) | 0.98  (0.92, 1.12) | 0.52  (0.46, 0.59) | 0.13  (0.12, 0.15) |
| **Vaccine Characteristic and Coverage Scenarios** | | | | | |
| 70% Efficacy | 22.2  (19.6, 25.9) | 20.2  (18, 23.6) | 1.35  (1.25, 1.55) | 0.70  (0.60, 0.79) | 0.18  (0.16, 0.21) |
| 5 years protection | 8.1  (6.6, 9.7) | 7.5  (6.1, 9) | 0.58  (0.54, 0.65) | 0.30  (0.26, 0.34) | 0.08  (0.07, 0.09) |
| 15 years protection | 17.1  (15.1, 20.4) | 15.5  (13.7, 18.6) | 1.01  (0.93, 1.14) | 0.53  (0.44, 0.58) | 0.14  (0.12, 0.16) |
| 20 years protection | 19.5  (17.2, 23.1) | 17.5  (15.6, 21) | 1.10  (1.01, 1.25) | 0.57  (0.48, 0.64) | 0.15  (0.13, 0.17) |
| Prevention of infection and disease | 21.9  (19.6, 25.3) | 19.8  (17.8, 23) | 1.3  (1.21, 1.5) | 0.67  (0.58, 0.76) | 0.17  (0.16, 0.2) |
| Efficacious with any infection status at vaccination | 16.4  (14.4, 19.2) | 15  (13.3, 17.6) | 1.01  (0.94, 1.16) | 0.53  (0.45, 0.59) | 0.14  (0.12, 0.16) |
| 2029 introduction | 13.5  (11.9, 15.9) | 11.9  (10.5, 14.2) | 0.67  (0.62, 0.76) | 0.34  (0.28, 0.38) | 0.09  (0.08, 0.10) |
| Lower coverage | 12.5  (10.7, 14.9) | 11.3  (9.8, 13.6) | 0.78  (0.72, 0.87) | 0.40  (0.34, 0.45) | 0.10  (0.09, 0.12) |
| Higher coverage | 14.9  (12.8, 17.7) | 13.6  (11.7, 16.2) | 0.94  (0.87, 1.06) | 0.49  (0.42, 0.54) | 0.13  (0.11, 0.15) |

*Abbreviations: IRR = incidence rate reduction, MRR = mortality rate reduction.*

### 9. Economic results

#### 9.1 M72/AS01_E_ scenarios

**Table S9.1** Incremental DALYs averted, incremental costs averted, and ICERs from the health-system

and societal perspectives for the M72/AS01_E_ *Vaccine Characteristic and Coverage Scenarios* compared to the no

new-vaccine baseline

| **Scenario** | **Incremental DALYs averted between 2025–2050**  **(millions)** | **Health-system perspective** | | **Societal perspective** | |
| --- | --- | --- | --- | --- | --- |
|  |  | **Incremental costs between 2025–2050 ($, millions)** | **ICERs  ($/DALY averted)** | **Incremental costs between 2025–2050 ($, millions)** | **ICERs  ($/DALY averted)** |
| Basecase | 3.7  (3.1, 4.1) | 255  (135, 399) | 69  (38, 110) | 64  (-59, 228) | 18  (cost-saving, 60) |
| **Vaccine Characteristic and Coverage Scenarios** | | | | | |
| 60% efficacy | 4.3  (3.7, 4.8) | 243  (115, 386) | 56  (28, 92) | 14  (-113, 190) | 3  (cost-saving, 42) |
| 70% efficacy | 4.9  (4.2, 5.5) | 232  (98, 378) | 47  (21, 77) | -34  (-172, 146) | cost-saving (-36, 28) |
| Lower efficacy in PLHIV | 3.4  (2.9, 3.8) | 254  (137, 400) | 74  (41, 118) | 78  (-46, 239) | 23  (cost-saving, 68) |
| Same efficacy in PLHIV | 4.0  (3.4, 4.5) | 256  (132, 398) | 64  (35, 102) | 44  (-79, 215) | 11  (cost-saving, 51) |
| 5 years protection | 2.8  (2.3, 3.1) | 276  (165, 419) | 100  (61, 156) | 141  (22, 297) | 51  (9, 108) |
| 15 years protection | 4.1  (3.5, 4.6) | 245  (119, 387) | 60  (31, 98) | 30  (-98, 201) | 7  (cost-saving, 47) |
| 20 years protection | 4.3  (3.7, 4.8) | 239  (111, 383) | 55  (27, 91) | 10  (-118, 186) | 2  (cost-saving, 41) |
| Prevention of infection and disease | 5.1  (4.3, 5.7) | 233  (98, 381) | 45  (21, 75) | -43  (-182, 139) | cost-saving  (cost-saving, 26) |
| Efficacious with current infection at vaccination | 2.3  (1.9, 2.6) | 289  (179, 434) | 127  (81, 199) | 180  (70, 342) | 80  (30, 150) |
| 2034 introduction | 2.4  (2.1, 2.7) | 214  (119, 337) | 88  (50, 141) | 94  (-10, 227) | 39  (cost-saving, 93) |
| Lower coverage | 2.9  (2.5, 3.3) | 193  (100, 305) | 66  (36, 106) | 42  (-53, 168) | 14  (cost-saving, 57) |
| Higher coverage | 4.4  (3.7, 4.9) | 318  (172, 496) | 72  (40, 114) | 91  (-61, 292) | 21  (cost-saving, 64) |

*Abbreviations: DALYs = disability-adjusted life years, ICERs = incremental cost-effectiveness ratio, US$ = United States Dollar. Values in cells are the mean and 95% uncertainty ranges.*

**Table S9.2** Total vaccination costs, and incremental diagnostic, treatment, and net costs between 2025–2050 for the M72/AS01_E_ scenarios from the health-system perspective. All costs are in US$ millions

| **Scenario** | **Vaccination costs** | **DS-TB diagnostic costs** | **RR-TB diagnostic costs** | **DS-TB treatment costs** | **RR-TB treatment costs** | **ART costs** | **Incremental cost** |
| --- | --- | --- | --- | --- | --- | --- | --- |
| Basecase | 333  (226, 489) | -3  (-6, -1) | -0.31  (-0.36, -0.26) | -84  (-147, -40) | -88  (-113, -68) | 97  (75, 123) | 255  (135, 399) |
| **Policy Scenarios** | | | | | | | |
| Younger ages (routine age 10, campaign for ages 11-34) | 405  (274, 594) | -3  (-6, -1) | -0.32  (-0.41, -0.30) | -97  (-167, -47) | -101  (-129, -79) | 150  (114, 196) | 354  (208, 528) |
| Older ages (campaign for ages 18-55) | 391  (265, 574) | -3  (-6, -1) | -0.33  (-0.39, -0.28) | -91  (-157, -43) | -96  (-123, -74) | 103  (78, 131) | 30  (166, 472) |
| **Vaccine Characteristic and Coverage Scenarios** | | | | | | | |
| 60% efficacy | 333  (226, 489) | -3  (-7, -1) | -0.36  (-0.43, -0.31) | -99  (-173, -47) | -104  (-133, -80) | 116  (89, 148) | 243  (115, 386) |
| 70% efficacy | 333  (226, 489) | -3  (-8, -1) | -0.41  (-0.49, -0.35) | -114  (-198, -54) | -119  (-153, -92) | 135  (104, 172) | 232 (98, 378) |
| Lower efficacy in PLHIV | 333  (226, 489) | -2  (-5, -1) | -0.29  (-0.34, -0.24) | -79  (-137, -38) | -83  (-105, -64) | 85  (65, 108) | 254  (137, 400) |
| Same efficacy in PLHIV | 333  (226, 489) | -3  (-6, -1) | -0.34  (-0.40, -0.28) | -92  (-162, -44) | -97  (-123, -75) | 115  (89, 147) | 256  (132, 398) |
| 5 years protection | 333  (226, 489) | -2  (-4, 0) | -0.23  (-0.27, -0.19) | -63  (-110, -30) | -66  (-84, -51) | 75  (57, 95) | 276  (165, 419) |
| 15 years protection | 333  (226, 489) | -3  (-6, -1) | -0.34  (-0.41, -0.29) | -94  (-164, -45) | -99  (-126, -76) | 107  (82, 136) | 245  (119, 387) |
| 20 years protection | 333  (226, 489) | -3  (-7, -1) | -0.36  (-0.43, -0.31) | -99  (-173, -47) | -104  (-133, -80) | 114  (87, 143) | 239  (111, 383) |
| Prevention of infection and disease | 333  (226, 489) | -4  (-8, -1) | -0.43  (-0.51, -0.36) | -117  (-205, -56) | -123  (-157, -95) | 144  (111, 183) | 233  (98, 381) |
| Efficacy with current infection at vaccination | 333  (226, 488) | -2  (-3, 0) | -0.19  (-0.23, -0.16) | -53  (-93, -25) | -55  -71, -43) | 65  (49, 84) | 289  (179, 434) |
| 2034 introduction | 283  (192, 415) | -2  (-4, 0) | -0.20  (-0.24, -0.17) | -56  (-98, -26) | -58  (-75, -45) | 47  (36, 60) | 214  (119, 337) |
| Lower coverage | 258  (175, 377) | -2  (-4, -1) | -0.24  (-0.29, -0.21) | -6  (-117, -32) | -70  (-89, -54) | 75  (57, 95) | 193  (100, 305) |
| Higher coverage | 409  (277, 600) | -3  (-7, -1) | -0.37  (-0.44, -0.31) | -101  (-175, -48) | -106  (-135, -82) | 119  (91, 151) | 318  (172, 496) |

*Abbreviations: DS-TB = drug-susceptible tuberculosis, RR-TB = rifampicin resistant tuberculosis, US$ = United States Dollars. Values in cells are the mean and 95% uncertainty ranges.*

**Table S9.3** Total vaccination costs, and incremental diagnostic, treatment, and net costs between 2025–2050 for the M72/AS01_E_ scenarios from the societal perspective. All costs in US$ millions.

| **Scenario** | **Vaccination costs** | **TB costs (diagnosis + treat)** | **ART costs** | **Non-medical costs** | **Indirect costs** | **Incremental cost** |
| --- | --- | --- | --- | --- | --- | --- |
| Basecase | 360  (236, 528) | -175  (-245, -125) | 97  (75, 123) | -78  (-92, -65) | -139  (-165, -116) | 64  (-59, 228) |
| **Policy Scenarios** | | | | | | |
| Younger ages (routine age 10, campaign for ages 11-34) | 423  (278, 620) | -190  (-266, -134) | 102  (78, 131) | -84  (-101, -71) | -151  (-179, -126) | 100  (-42, 290) |
| Older ages (campaign for ages 18-55) | 438  (288, 642) | -201  (-281, -145) | 149  (114, 195) | -89  (-105, -75) | -160  (-188, -134) | 137  (-10, 327) |
| **Vaccine Characteristic and Coverage Scenarios** | | | | | | |
| 60% efficacy | 360  (236, 528) | -207  (-289, -147) | 116  (89, 147) | -92  (-109, -76) | -164  (-195, -136) | 14  (-113, 190) |
| 70% efficacy | 360  (236, 528) | -237  (-330, -168) | 135  (104, 172) | -105  (-125, -87) | -188  (-223, -156) | -34  (-172, 146) |
| Lower efficacy in PLHIV | 360  (236, 528) | -164  (-229, -116) | 85  (66, 108) | -73  (-86, -60) | -130  (-154, -108) | 78  (-46, 239) |
| Same efficacy in PLHIV | 360  (236, 528) | -193  (-269, -138) | 114  (89, 146) | -85  (-101, -71) | -153  (-181, -127) | 44  (-79, 215) |
| 5 years protection | 360  (236, 528) | -131  (-184, -93) | 74  (58, 95) | -58  (-69, -48) | -104  (-123, -87) | 141  (22, 297) |
| 15 years protection | 360  (236, 528) | -195  (-273, -139) | 107  (82, 136) | -87  (-103, -72) | -155  (-184, -129) | 30  (-98, 201) |
| 20 years protection | 360  (236, 528) | -207  (-289, -147) | 112  (87, 143) | -92  (-109, -76) | -164  (-195, -136) | 10  (-118, 186) |
| Prevention of infection and disease | 360  (236, 528) | -245  (-340, -175) | 144  (111, 182) | -108  (-129, -90) | -194  (-230, -161) | -43  (-182, 139) |
| Efficacy with current infection at vaccination | 360  (237, 528) | -110  (-157, -79) | 65  (50, 84) | -49  (-58, -40) | -87  (-104, -72) | 180  (70, 342) |
| 2036 introduction | 306  (201, 448) | -116  (-162, -83) | 47  (36, 60) | -51  (-61, -42) | -92  (-110, -76) | 94  (-10, 227) |
| Lower coverage | 279  (183, 408) | -139  (-194, -99) | 74  (58, 94) | -62  (-73, -51) | -110  (-131, -92) | 42  (-53, 168) |
| Higher coverage | 442  (290, 649) | -210  (-293, -149) | 118  (91, 151) | -93  (-111, -77) | -167  (-198, -139) | 91  (-61, 292) |

*Abbreviations: DS-TB = drug-susceptible tuberculosis, RR-TB = rifampicin resistant tuberculosis, US$ = United States Dollars. Values in cells are the mean and 95% uncertainty ranges.*

#### 9.2 BCG-revaccination scenarios

**Table S9.4** Incremental DALYs averted, incremental costs averted, and ICERs from the health-system

and societal perspectives for BCG-revaccination *Vaccine Characteristic and Coverage Scenarios* compared to

the no-new-vaccine baseline

| **Scenario** | **Incremental DALYs averted between 2025–2050**  **(millions)** | **Health-system perspective** | | **Societal perspective** | |
| --- | --- | --- | --- | --- | --- |
|  |  | **Incremental costs between 2025–2050 ($, millions)** | **ICERs  ($/DALY averted)** | **Incremental costs between 2025–2050 ($, millions)** | **ICERs  ($/DALY averted)** |
| Basecase | 2.2  (1.9, 2.4) | 50  (-11, 118) | 23  (cost-saving, 54) | -57  (-118, 27) | cost-saving  (cost-saving, 11) |
| **Vaccine Characteristic and Coverage Scenarios** | | | | | |
| 70% Efficacy | 3.4  (3, 3.8) | 28  (-49, 103) | 8  (cost-saving, 30) | -149  (-222, -53) | cost-saving  (cost-saving, -15) |
| 5 years protection | 1.5  (1.3, 1.7) | 65  (10, 133) | 43  (7, 90) | -5  (-62, 74) | cost-saving  (cost-saving, 48) |
| 15 years protection | 2.5  (2.2, 2.8) | 42  (-22, 113) | 17  (cost-saving, 44) | -85  (-149, 3) | cost-saving  (cost-saving, 1) |
| 20 years protection | 2.7  (2.4, 3) | 37  (-30, 109) | 14  (cost-saving, 39) | -102  (-170, -13) | cost-saving |
| Prevention of infection and disease | 3.3  (2.9, 3.6) | 29  (-46, 102) | 9  (cost-saving, 31) | -140  (-211, -47) | cost-saving |
| Efficacy with any infection status at vaccination | 2.6  (2.3, 2.9) | 45  (-21, 116) | 17  (cost-saving, 44) | -86  (-149, 2) | cost-saving  (cost-saving, 1) |
| 2029 introduction | 1.5  (1.4, 1.7) | 38  (-10, 94) | 24  (cost-saving, 62) | -37  (-88, 29) | cost-saving  (cost-saving, 18) |
| Lower coverage | 2  (1.7, 2.2) | 44  (-11, 103) | 22  (cost-saving, 52) | -53  (-107, 22) | cost-saving  (cost-saving, 10) |
| Higher coverage | 2.4  (2.1, 2.6) | 57  (-10, 133) | 24  (cost-saving, 56) | -60  (-127, 33) | cost-saving  (cost-saving, 13) |

*Abbreviations: DALYs = disability-adjusted life years, ICERs = incremental cost-effectiveness ratio, US$ = United States Dollar. Values in cells are the mean and 95% uncertainty ranges.*

**Table S9.5** Total vaccination costs, and incremental diagnostic, treatment, and net costs between 2025–2050 for the BCG-revaccination scenarios from the health-system perspective. All costs are in US$ millions

| **Scenario** | **Vaccination costs** | **DS-TB diagnostic costs** | **RR-TB diagnostic costs** | **DS-TB treatment costs** | **RR-TB treatment costs** | **ART costs** | **Incremental cost** |
| --- | --- | --- | --- | --- | --- | --- | --- |
| Basecase | 91  (39, 166) | -1  (-3, 0) | -0.17  (-0.20, -0.14) | -47  (-82, -22) | -49  (-63, -37) | 57  (43, 72) | 50  (-11, 118) |
| **Policy Scenarios** | | | | | | | |
| Older ages (routine age 15, campaign for ages 16-34) | 130  (55, 237) | -1  (-3, 0) | -0.18  (-0.21, -0.15) | -49  (-88, -23) | -52  (-67, -39) | 67  (52, 84) | 93  (15, 192) |
| Basecase plus on ART ages 11-18 | 92  (40, 168) | -1  (-3, 0) | -0.18  (-0.21, -0.15) | -49  (-85, -23) | -51  (-65, -39) | 62  (47, 78) | 52  (-11, 121) |
| Basecase plus on ART ages 10+ | 123  (52, 228) | -2  (-4, 0) | -0.21  (-0.24, -0.17) | -56  (-98, -27) | -59  (-76, -45) | 82  (62, 107) | 88  (8, 183) |
| Basecase plus virally suppressed ages 10+ | 122  (52, 226) | -2  (-4, 0) | -0.20  (-0.24, -0.17) | -56  (-97, -27) | -59  (-76, -45) | 81  (61, 105) | 86  (7, 181) |
| **Vaccine Characteristic and Coverage Scenarios** | | | | | | | |
| 70% Efficacy | 92  (39, 167) | -2  (-5, -1) | -0.27  (-0.32, -0.23) | -74  (-128, -35) | -77  (-99, -59) | 89  (68, 113) | 28  (-49, 103) |
| 5 years protection | 92  (39, 167) | -1  (-2, 0) | -0.12  (-0.14, -0.10) | -32  (-56, -15) | -34  (-43, -26) | 40  (31, 51) | 65  (10, 133) |
| 15 years protection | 91  (39, 166) | -2  (-4, 0) | -0.20  (-0.23, -0.17) | -54  (-95, -26) | -57  (-73, -44) | 65  (49, 82) | 42  (-22, 113) |
| 20 years protection | 91  (39, 166) | -2  (-4, 0) | -0.22  (-0.26, -0.18) | -59  (-104, -28) | -62  (-79, -47) | 70  (53, 88) | 37  (-30, 109) |
| Prevention of infection and disease | 91  (39, 166) | -2  (-5, -1) | -0.26  (-0.30, -0.23) | -70  (-122, -33) | -74  (-94, -56) | 84  (64, 107) | 29  (-46, 102) |
| Efficacy with any infection status at vaccination | 92  (40, 168) | -2  (-4, 0) | -0.20  (-0.24, -0.17) | -56  (-97, -26) | -59  (-74, -45) | 69  (52, 87) | 45  (-21, 116) |
| 2029 introduction | 77  (33, 139) | -1  (-2, 0) | -0.12  (-0.14, -0.10) | -33  (-58, -16) | -35  (-45, -27) | 31  (23, 39) | 38  (-10, 94) |
| Lower coverage | 81  (35, 147) | -1  (-3, 0) | -0.15  (-0.18, -0.13) | -42  (-73, -20) | -44  (-56, -34) | 51  (39, 64) | 44  (-11, 103) |
| Higher coverage | 102  (44, 185) | -2  (-3, 0) | -0.19  (-0.22, -0.16) | -51  (-89, -24) | -54  (-69, -41) | 62  (47, 79) | 57  (-10, 133) |

*Abbreviations: DS-TB = drug-susceptible tuberculosis, RR-TB = rifampicin resistant tuberculosis, US$ = United States Dollars. Values in cells are the mean and 95% uncertainty ranges.*

**Table S9.6** Total vaccination costs, and incremental diagnostic, treatment, and net costs between 2025–2050

for the BCG-revaccination scenarios from the societal perspective. All costs in US$ millions

| **Scenario** | **Vaccination costs** | **TB costs (diagnosis + treat)** | **ART costs** | **Non-medical costs** | **Indirect costs** | **Incremental cost** |
| --- | --- | --- | --- | --- | --- | --- |
| Basecase | 104  (44, 184) | -97  (-136, -69) | 56  (43, 72) | -43  (-52, -36) | -77  (-92, -64) | -57  (-118, 27) |
| **Policy Scenarios** | | | | | | |
| Older ages (routine age 15, campaign for ages 16-34) | 148  (62, 264) | -103  (-148, -72) | 67  (52, 85) | -46  (-55, -36) | -82  (-97, -65) | -15  (-102, 101) |
| Basecase plus on ART ages 11-18 | 105  (44, 185) | -101  (-141, -71) | 61  (46, 78) | -45  (-54, -37) | -80  (-96, -66) | -60  (-121, 26) |
| Basecase plus on ART ages 10+ | 141  (58, 250) | -117  (-164, -82) | 81  (61, 107) | -52  (-63, -43) | -93  (-112, -77) | -40  (-122, 70) |
| Basecase plus virally suppressed ages 10+ | 139  (58, 247) | -116  (-163, -82) | 80  (60, 105) | -52  (-62, -43) | -92  (-111, -76) | -40  (-121, 68) |
| **Vaccine Characteristic and Coverage Scenarios** | | | | | | |
| 70% Efficacy | 105  (44, 185) | -15  (-215, -108) | 89  (68, 113) | -68  (-82, -56) | -121  (-146, -101) | -149  (-222, -53) |
| 5 years protection | 105  (44, 184) | -67  (-93, -47) | 40  (30, 51) | -30  (-35, -24) | -53  (-63, -44) | -5  (-62, 74) |
| 15 years protection | 10  (44, 183) | -113  (-158, -80) | 64  (49, 82) | -50  (-60, -41) | -90  (-108, -74) | -85  (-149, 3) |
| 20 years protection | 104  (44, 183) | -12  (-173, -87) | 69  (53, 89) | -55  (-65, -45) | -98  (-116, -80) | -102  (-170, -13) |
| Prevention of infection and disease | 104  (44, 184) | -146  (-206, -103) | 83  (63, 107) | -65  (-78, -54) | -116  (-139, -97) | -140  (-211, -47) |
| Efficacy with any infection status at vaccination | 105  (44, 186) | -116  (-162, -82) | 68  (53, 88) | -51  (-61, -43) | -92  (-109, -77) | -86  (-149, 2) |
| 2029 introduction | 87  (37, 153) | -69  (-97, -49) | 30  (23, 39) | -31  (-37, -25) | -55  (-66, -45) | -37  (-88, 29) |
| Lower coverage | 92  (39, 162) | -88  (-122, -62) | 50  (38, 64) | -39  (-46, -32) | -69  (-83, -57) | -53  (-107, 22) |
| Higher coverage | 116  (49, 205) | -107  (-148, -75) | 62  (47, 79) | -47  (-56, -39) | -85  (-101, -70) | -60  (-127, 33) |

*Abbreviations: DS-TB = drug-susceptible tuberculosis, RR-TB = rifampicin resistant tuberculosis, US$ = United States Dollars. Values in cells are the mean and 95% uncertainty ranges.*

### 10. References

1.	Clark, R.A., et al., *The impact of alternative delivery strategies for novel tuberculosis vaccines in low-income and middle-income countries: a modelling study.* Lancet Glob Health, 2023. **11**(4): p. e546-e555.

2.	Clark, R.A., et al., *New tuberculosis vaccines in India: Modelling the potential health and economic impacts of adolescent/adult vaccination with M72/AS01 (E) and BCG-revaccination.* medRxiv, 2023.

3.	United Nations Department of Economic and Social Affairs, P.D. *World Population Projections (2019 revision)*. 2019; Available from: <https://population.un.org/wpp/Download/Standard/Population/>.

4.	Tiemersma, E.W., et al., *Natural history of tuberculosis: duration and fatality of untreated pulmonary tuberculosis in HIV negative patients: a systematic review.* PLoS One, 2011. **6**(4): p. e17601.

5.	Quaife, M., et al., *Post-tuberculosis mortality and morbidity: valuing the hidden epidemic.* Lancet Respir Med, 2020. **8**(4): p. 332-333.

6.	Emery, J.C., et al., *Estimating the contribution of subclinical tuberculosis disease to transmission – an individual patient data analysis from prevalence surveys.* 2022: p. 2022.06.09.22276188.

7.	Emery, J.C., et al., *Self-clearance of Mycobacterium tuberculosis infection: implications for lifetime risk and population at-risk of tuberculosis disease.* Proc Biol Sci, 2021. **288**(1943): p. 20201635.

8.	Dye, C. and B.G. Williams, *Eliminating human tuberculosis in the twenty-first century.* J R Soc Interface, 2008. **5**(23): p. 653-62.

9.	Abu-Raddad, J.L., et al., *Epidemiological benefits of more-effective tuberculosis vaccines, drugs and diagnostics.* PNAS, 2009. **106**(33).

10.	Marx, F.M., et al., *The temporal dynamics of relapse and reinfection tuberculosis after successful treatment: a retrospective cohort study.* Clin Infect Dis, 2014. **58**(12): p. 1676-83.

11.	Gomes, M.G., L.J. White, and G.F. Medley, *The reinfection threshold.* J Theor Biol, 2005. **236**(1): p. 111-3.

12.	Sutherland, I., E. Svandova, and S. Radhakrishna, *The development of clinical tuberculosis following infection with tubercle bacilli.* Tubercle, 1982. **62**(4): p. 255-68.

13.	Vynnycky, E. and P.E.M. Fine, *The natural history of tuberculosis: the implications of age-dependent risks of disease and the role of reinfection.* Epidemiology and Infection, 1997. **119**: p. 183-201.

14.	Stover, J., R. McKinnon, and B. Winfrey, *Spectrum: a model platform for linking maternal and child survival interventions with AIDS, family planning and demographic projections.* Int J Epidemiol, 2010. **39 Suppl 1**: p. i7-10.

15.	Mahy, M., et al., *Derivation of parameters used in Spectrum for eligibility for antiretroviral therapy and survival on antiretroviral therapy.* Sex Transm Infect, 2010. **86 Suppl 2**: p. ii28-34.

16.	eligibility for, A.R.T.i.l.i.c.c., et al., *Duration from seroconversion to eligibility for antiretroviral therapy and from ART eligibility to death in adult HIV-infected patients from low and middle-income countries: collaborative analysis of prospective studies.* Sex Transm Infect, 2008. **84 Suppl 1**(Suppl_1): p. i31-i36.

17.	Granich, R.M., et al., *Universal voluntary HIV testing with immediate antiretroviral therapy as a strategy for elimination of HIV transmission: a mathematical model.* Lancet, 2009. **373**(9657): p. 48-57.

18.	Cori, A., et al., *HPTN 071 (PopART): a cluster-randomized trial of the population impact of an HIV combination prevention intervention including universal testing and treatment: mathematical model.* PLoS One, 2014. **9**(1): p. e84511.

19.	Mberi, M.N., et al., *Determinants of loss to follow-up in patients on antiretroviral treatment, South Africa, 2004-2012: a cohort study.* BMC Health Serv Res, 2015. **15**: p. 259.

20.	Van Cutsem, G., et al., *Correcting for mortality among patients lost to follow up on antiretroviral therapy in South Africa: a cohort analysis.* PLoS One, 2011. **6**(2): p. e14684.

21.	Fenner, L., et al., *Early mortality and loss to follow-up in HIV-infected children starting antiretroviral therapy in Southern Africa.* J Acquir Immune Defic Syndr, 2010. **54**(5): p. 524-32.

22.	Sengayi, M., et al., *Predictors of loss to follow-up among children in the first and second years of antiretroviral treatment in Johannesburg, South Africa.* Glob Health Action, 2013. **6**: p. 19248.

23.	Chandiwana, N., et al., *High loss to follow-up of children on antiretroviral treatment in a primary care HIV clinic in Johannesburg, South Africa.* Medicine (Baltimore), 2018. **97**(29): p. e10901.

24.	Fatti, G., et al., *Antiretroviral treatment outcomes amongst older adults in a large multicentre cohort in South Africa.* PLoS One, 2014. **9**(6): p. e100273.

25.	Stover, J., et al., *Updates to the Spectrum/AIM model for estimating key HIV indicators at national and subnational levels.* AIDS, 2019. **33 Suppl 3**(Suppl 3): p. S227-S234.

26.	Johansson, K.A., B. Robberstad, and O.F. Norheim, *Further benefits by early start of HIV treatment in low income countries: survival estimates of early versus deferred antiretroviral therapy.* AIDS Res Ther, 2010. **7**(1): p. 3.

27.	Bruchfeld, J., M. Correia-Neves, and G. Kallenius, *Tuberculosis and HIV Coinfection.* Cold Spring Harb Perspect Med, 2015. **5**(7): p. a017871.

28.	Selwyn, P.A., et al., *A prospective study of the risk of tuberculosis among intravenous drug users with human immunodeficiency virus infection.* N Engl J Med, 1989. **320**(9): p. 545-50.

29.	Kwan, C.K. and J.D. Ernst, *HIV and tuberculosis: a deadly human syndemic.* Clin Microbiol Rev, 2011. **24**(2): p. 351-76.

30.	Mukadi, Y.D., D. Maher, and A. Harries, *Tuberculosis case fatality rates in high HIV prevalence populations in sub-Saharan Africa.* AIDS, 2001. **15**(2): p. 143-52.

31.	Sonnenberg, P., et al., *How soon after infection with HIV does the risk of tuberculosis start to increase? A retrospective cohort study in South African gold miners.* J Infect Dis, 2005. **191**(2): p. 150-8.

32.	Lawn, S.D., et al., *Impact of HIV infection on the epidemiology of tuberculosis in a peri-urban community in South Africa: the need for age-specific interventions.* Clin Infect Dis, 2006. **42**(7): p. 1040-7.

33.	Lawn, S.D., et al., *Short-term and long-term risk of tuberculosis associated with CD4 cell recovery during antiretroviral therapy in South Africa.* AIDS, 2009. **23**(13): p. 1717-25.

34.	Badri, M., D. Wilson, and R. Wood, *Effect of highly active antiretroviral therapy on incidence of tuberculosis in South Africa: a cohort study.* Lancet, 2002. **359**(9323): p. 2059-64.

35.	Suthar, A., et al., *Antiretroviral therapy for prevention of tuberculosis in adults with HIV: a systematic review and meta-analysis.* Plos Medicine, 2012. **9**(7).

36.	Ackah, A.N., et al., *Response to treatment, mortality, and CD4 lymphocyte counts in HIV-infected persons with tuberculosis in Abidjan, Cote d'Ivoire.* Lancet, 1995. **345**(8950): p. 607-10.

37.	Corbett, E.L., et al., *Stable incidence rates of tuberculosis (TB) among Human Immunodeficiency Virus (HIV)-negative South African gold miners during a decade of epidemic HIV-associated TB.* JID, 2003. **188**.

38.	World Health Organisation. *WHO Treatment Outcomes Database*. 2017 [accessed 2017 20th September]; Available from: <https://extranet.who.int/tme/generateCSV.asp?ds=outcomes>.

39.	R Core Team, *R: A language and environment for statistical computing*, R Foundation for Statistical computing, Editor. 2013: Vienna.

40.	Andrianakis, I., et al., *History matching of a complex epidemiological model of human immunodeficiency virus transmission by using variance emulation.* J R Stat Soc Ser C Appl Stat, 2017. **66**(4): p. 717-740.

41.	Andrianakis, I., et al., *Bayesian history matching of complex infectious disease models using emulation: a tutorial and a case study on HIV in Uganda.* PLoS Comput Biol, 2015. **11**(1): p. e1003968.

42.	Iskauskas, A., *hmer: History Matching and Emulation package*. 2022.

43.	World Health Organisation. *Global Tuberculosis Report*. 2022 [accessed 2021; Available from: <https://apps.who.int/iris/handle/10665/363752>.

44.	Moyo, S., et al., *Prevalence of bacteriologically confirmed pulmonary tuberculosis in South Africa, 2017-19: a multistage, cluster-based, cross-sectional survey.* Lancet Infect Dis, 2022. **22**(8): p. 1172-1180.

45.	Yates, T.A., et al., *Socio-economic gradients in prevalent tuberculosis in Zambia and the Western Cape of South Africa.* Trop Med Int Health, 2018. **23**(4): p. 375-390.

46.	Harling, G., R. Ehrlich, and L. Myer, *The social epidemiology of tuberculosis in South Africa: a multilevel analysis.* Soc Sci Med, 2008. **66**(2): p. 492-505.

47.	UN AIDS, *HIV estimates with uncertianty bounds 1990-2020*. 2021.

48.	Nemes, E., et al., *Prevention of M. tuberculosis Infection with H4:IC31 Vaccine or BCG Revaccination.* N Engl J Med, 2018. **379**(2): p. 138-149.

49.	Tait, D.R., et al., *Final Analysis of a Trial of M72/AS01(E) Vaccine to Prevent Tuberculosis.* N Engl J Med, 2019. **381**(25): p. 2429-2439.

50.	World Health Organisation. *WHO preferred product characteristics for new tuberculosis vaccines*. 2018; Available from: <http://apps.who.int/iris/handle/10665/273089>.

51.	Pelzer, P.T., et al., *Potential implementation strategies, acceptability, and feasibility of new and repurposed TB vaccines.* PLOS Glob Public Health, 2022. **2**(5): p. e0000076.

52.	Kumarasamy, N., et al., *Long-term safety and immunogenicity of the M72/AS01E candidate tuberculosis vaccine in HIV-positive and -negative Indian adults: Results from a phase II randomized controlled trial.* Medicine (Baltimore), 2018. **97**(45): p. e13120.

53.	World Health Organisation, *Revised BCG vaccination guidelines for infants at risk for HIV infection.* Weekly Epidemiological Record, 2007. **82**(21): p. 193-196.

54.	World Health, O., *BCG vaccine: WHO position paper, February 2018 – Recommendations.* Vaccine, 2018. **36**(24): p. 3408-3410.

55.	GBD 2019 Disease and Injuries Collaborators, *Global burden of 369 diseases and injuries in 204 countries and territories, 1990-2019: a systematic analysis for the Global Burden of Disease Study 2019.* The Lancet, 2020. **396**(10258): p. 1204-1222.

56.	UNICEF. *Bacillus Calmette–Guérin (BCG) vaccine price data*. 2021; Available from: <https://www.unicef.org/supply/documents/bacillus-calmettegu%C3%A9rin-bcg-vaccine-price-data>.

57.	Gavi The Vaccine Alliance. *GAVI Alliance Vaccine Introduction Grant and Operational Support for Campaigns Policy. Version 1.0*. 2013; Available from: <www.gavi.org>

58.	UNICEF. *Costs of vaccinating a child*. 2020; Available from: <https://immunizationeconomics.org/recent-activity/2021/6/15/standard-costs-of-vaccinating-a-child>

59.	Prosser, L.A., et al., *Non-traditional settings for influenza vaccination of adults: costs and cost effectiveness.* Pharmacoeconomics, 2008. **26**(2): p. 163-78.

60.	The World Bank. *World development indicators*. 2022; Available from: <https://data.worldbank.org/>.

61.	Ochalek, J., J. Lomas, and K. Claxton, *Estimating health opportunity costs in low-income and middle-income countries: a novel approach and evidence from cross-country data.* BMJ Glob Health, 2018. **3**(6): p. e000964.

62.	Jha, S., et al., *Cost-Effectiveness of Automated Digital Microscopy for Diagnosis of Active Tuberculosis.* PLoS One, 2016. **11**(6): p. e0157554.

63.	Siapka, M., et al., *Cost of tuberculosis treatment in low- and middle-income countries: systematic review and meta-regression.* Int J Tuberc Lung Dis, 2020. **24**(8): p. 802-810.

64.	van Rensburg, C., et al., *Cost outcome analysis of decentralized care for drug-resistant tuberculosis in Johannesburg, South Africa.* PLoS One, 2019. **14**(6): p. e0217820.

65.	Masuku, S., *The cost and impact of MDR-TB treatment guideline changes in South Africa*, in *CROI*. 2019: Seattle, Washington.

66.	Foster, N., et al., *Strengthening health systems to improve the value of tuberculosis diagnostics in South Africa: A cost and cost-effectiveness analysis.* PLoS One, 2021. **16**(5): p. e0251547.

67.	Meyer-Rath, G., et al., *The per-patient costs of HIV services in South Africa: Systematic review and application in the South African HIV Investment Case.* PLoS One, 2019. **14**(2): p. e0210497.
